# Influenza A H1N1–mediated pre-existing immunity to SARS-CoV-2 predicts COVID-19 outbreak dynamics

**DOI:** 10.1101/2021.12.23.21268321

**Authors:** Nerea Martín Almazán, Afsar Rahbar, Marcus Carlsson, Tove Hoffman, Linda Kolstad, Bengt Rönnberg, Mattia Russel Pantalone, Ilona Lewensohn Fuchs, Anna Nauclér, Mats Ohlin, Mariusz Sacharczuk, Piotr Religa, Stefan Amér, Christian Molnár, Åke Lundkvist, Andres Susrud, Birger Sörensen, Cecilia Söderberg-Nauclér

**Affiliations:** Department of Medicine, Unit for Microbial Pathogenesis, Karolinska Institutet, Stockholm, Sweden; Department of Neurology, Karolinska University Hospital, Stockholm, Sweden; Department of Laboratory Medicine, Division of Pathology, Karolinska Institutet, Stockholm, Sweden; Centre for the Mathematical Sciences, Lund University; Zoonosis Science Center (ZSC), Department of Medical Biochemistry and Microbiology (IMBIM), Uppsala University, Uppsala, Sweden; Division of Clinical Microbiology, Department of Labortory Medicine, Karolinska Institutet, Stockholm, Sweden; Department of Clinical Microbiology, Karolinska University Hospital, Stockholm, Sweden; Department of Immunotechnology and SciLifeLab Human Antibody Therapeutics Infrastructure Unit, Lund University, Lund, Sweden; Faculty of Pharmacy with the Laboratory Medicine Division, Department of Pharmacodynamics, Medical University of Warsaw, Centre for Preclinical Research and Technology, Banacha 1B, Warsaw, Poland; Department of Experimental Genomics, Institute of Genetics and Animal Biotechnology, Polish Academy of Sciences, Postepu 36A, Jastrzebiec, Poland; Familjeläkarna Saltsjöbaden, Saltsjöbaden, Sweden; Department of Neurobiology, Care Sciences and Society, NVS, Karolinska Institutet, Stockholm, Sweden; Immunor AS, Oslo, Norway

**Keywords:** SARS-CoV-2, influenza A H1N1, HLA types, pre-existing immunity, molecular mimicry, mathematical models

## Abstract

**Background:** Susceptibility to SARS-CoV-2 infections is highly variable, ranging from asymptomatic and mild infections in most, to deadly outcome in few. This individual difference in susceptibility and outcome could be mediated by a cross protective pre-immunity, but the nature of this pre-immunity has remained elusive.

**Methods:** Antibody epitope sequence similarities and cross-reactive T cell peptides were searched for between SARS-CoV-2 and other pathogens. We established an ELISA test, a Luminex Multiplex bead array assay and a T cell assay to test for presence of identified peptide specific immunity in blood from SARS-CoV-2 positive and negative individuals. Mathematical modelling tested if SARS-CoV-2 outbreak dynamics could be predicted.

**Findings:** We found that peptide specific antibodies induced by influenza A H1N1 (flu) strains cross react with the most critical receptor binding motif of the SARS-CoV-2 spike protein that interacts with the ACE2 receptor. About 55–73% of COVID-19 negative blood donors in Stockholm had detectable antibodies to this peptide, NGVEGF, in the early pre-vaccination phase of the pandemic, and seasonal flu vaccination trended to enhance SARS-CoV-2 antibody and T cell immunity to this peptide. Twelve identified flu/SARS-CoV-2 cross-reactive T cell peptides could mediate protection against SARS-CoV-2 in 40–71% of individuals, depending on their HLA type. Mathematical modelling taking pre-immunity into account could fully predict pre-omicron SARS-CoV-2 outbreaks.

**Interpretation:** The presence of a specific cross-immunity between Influenza A H1N1 strains and SARS-CoV-2 provides mechanistic explanations to the epidemiological observations that influenza vaccination protects people against SARS-CoV-2 infection.

## Introduction

Although the COVID-19 epidemic was declared a pandemic in March 2020, it is still not fully under control, and emerging mutant strains continue to cause great concern. SARS-CoV-2 is considered a new virus to humans. Therefore, a major impact was expected. Mathematical modeling predicted an infection rate of at least 70% within a few months, suggesting catastrophic scenarios of collapsed health care systems and high death tolls if strict nonpharmacological mitigation strategies were not implemented (1). However, after the first wave, measured seroprevalence levels were less than 25% in a majority of hard-hit locations. In Stockholm, Sweden, a seroprevalence of 12% was reached in September 2020 after the first wave and under less strict nonpharmacological interventions than in most other western countries.

The SARS-CoV-2 infection affects people very differently. About 50% of infections caused by the original Wuhan strain were asymptomatic. Among people with symptomatic infections, 80% had mild symptoms; 20% developed severe disease and required hospital care, and 3–5% were admitted to the intensive care unit (2, 3). People over 70 years of age and those with obesity, type II diabetes, or hypertension are at higher risk of severe disease (4), and the impact on some patients, even young previously healthy people, can be disastrous. A possible explanation for differences in susceptibility is pre-existing protective immunity, suggested by the unexpected decline in infections during the first wave of the pandemic. In June 2020, 11% were estimated to be seropositive for SARS-CoV-2 in Stockholm, yet the decline started in early April, about 1 month after the onset of the first wave, despite rather limited mitigation strategies compared to other countries. From mid-March 2020, high schools and universities were on distance learning, people were expected to work from home and, if possible, avoid public transportation. Frequent hand washing, social distancing, and staying at home if feeling sick were the main recommendations from health authorities and the government. A lock-down was never implemented, elementary and middle schools remained open, and until January 2021, face masks were not recommended, even in the care of vulnerable patients in hospitals.

The decline of cases in larger cities in Sweden continued from April, which implied some kind of protective immunity among people. Infection rates on cruise ships during the first wave also peaked at ∼20%, even though many passengers were older than 70 (5, 6). By May 2020, 19.1% of 2149 staff members at Danderyd’s Hospital in Stockholm tested positive for SARS-CoV-2 antibodies (7). Furthermore, rarely more than 15–20% of household members became infected with pre-omicron strains by a family member with confirmed COVID-19 (8). At several elder care homes in Stockholm and Uppsala in Sweden, about 23% of personnel rapidly became antibody positive during the first wave (9). In New York, seroprevalence was 23.6% after the spring of 2020 (10). We hypothesized that this pattern of declining viral spread and the apparent protection of about 75-80% of the population from severe COVID-19 disease before omicron evolved is best explained by a pre-existing immunity, which would also contribute to herd immunity thresholds. To test this hypothesis, we used mathematical models to study the effects of factors such as nonpharmacological interventions, age, interactive patterns, mobility, and pre-immunity on viral outbreaks. It proved impossible to match modeled and real data without incorporating a pre-existing immunity level of 50–60% (11, 12). We therefore set out to identify the source of the pre-immunity.

Pre-immunity to SARS-CoV-2 is most likely mediated by previous infections. Indeed, 40–60% of healthy blood donors, including those giving blood before SARS-CoV-2 existed, respond to SARS-CoV-2 peptides in vitro (13-17), and pre-existing polymerase-peptide specific T cells expand, particularly in patients with abortive infections (16). Such T-cell pre-immunity was implied to be caused by common cold coronaviruses (18-20) and could contribute to herd immunity levels (21-23). Hence, in some people, T cells trained to recognize unrelated pathogen peptides may protect against severe COVID-19 through cross-immunity (or molecular mimicry). However, as this virus can be transmitted by aerosols (24), it is unlikely that T cells protect against infection with SARS-CoV-2 on a population level. Instead, antibody protection is expected to be required.

Neutralizing antibodies to SARS-CoV-2 have been found in pre-pandemic sera (25) and must therefore have been triggered by another pathogen, potentially by other common cold coronaviruses (19). One report suggested that 44% of children and 5.7% of adults have antibodies to common coronaviruses that can confer neutralizing activity against SARS-CoV-2 (20). However, although coronavirus antibody titers are boosted by SARS-CoV-2 infection, these are not considered protective (26). Furthermore, this level of pre-immunity would not explain the slow-down of SARS-CoV-2 spread when about 20% of a population becomes infected. We therefore searched for potential cross-reactivities between SARS-CoV-2 and other pathogens.

## Results

### Identification of potential cross-reactivities between SARS-CoV-2 and other pathogens

A BLAST search for cross-reactive protein sequences between SARS-CoV-2 and any other unrelated pathogen identified SARS-CoV (2004) with 76% homology, MERS-CoV (2012) with 35.1% homology and other pathogens such as non-human coronaviruses, human coronaviruses and of interest the cysteine bond of the SARS-CoV-2 homology to the neuraminidase protein of H1N1 Nagasaki and Kyoto strains. This led us to further analyze any potential homologies to other influenza strains which could possibly explain the SARS-CoV-2 pre-immunity. We next used a method focusing on small 6-mer peptides aiming to search for cross-reactive epitopes that are optimal for antibody binding, identified a peptide in SARS-CoV-2, NGVEGF (Fig. 1a), that is identical to a peptide in the neuraminidase of two strains of influenza A H1N1 (swine flu): Nagasaki/07N005/2008 and Kyoto/07K520/2008. NGVEGF was not found in any H1N1 strain sequenced before 2008 (Table S1). A variant, NGVKGF, was present in 99.3% of swine flu strains (*n* = 18,972) sequenced after 2008 and in 31.4% of strains (*n* = 1467) sequenced before 2008 (Table S1). The NGVKGF peptide is present in SARS-CoV-2 variants from Brazil (Gamma, P1), South Africa (Beta, B.1.351, V 501Y.V2), and New York (Iota, B.1.526), which carry an E484K mutation.

**Figure. 1.**
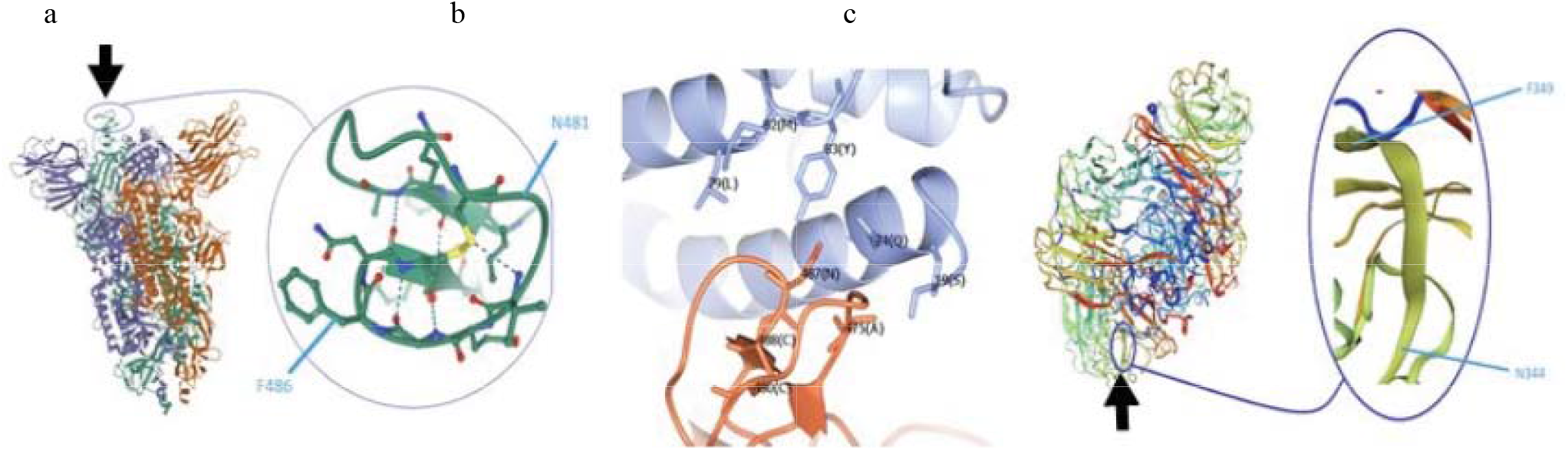
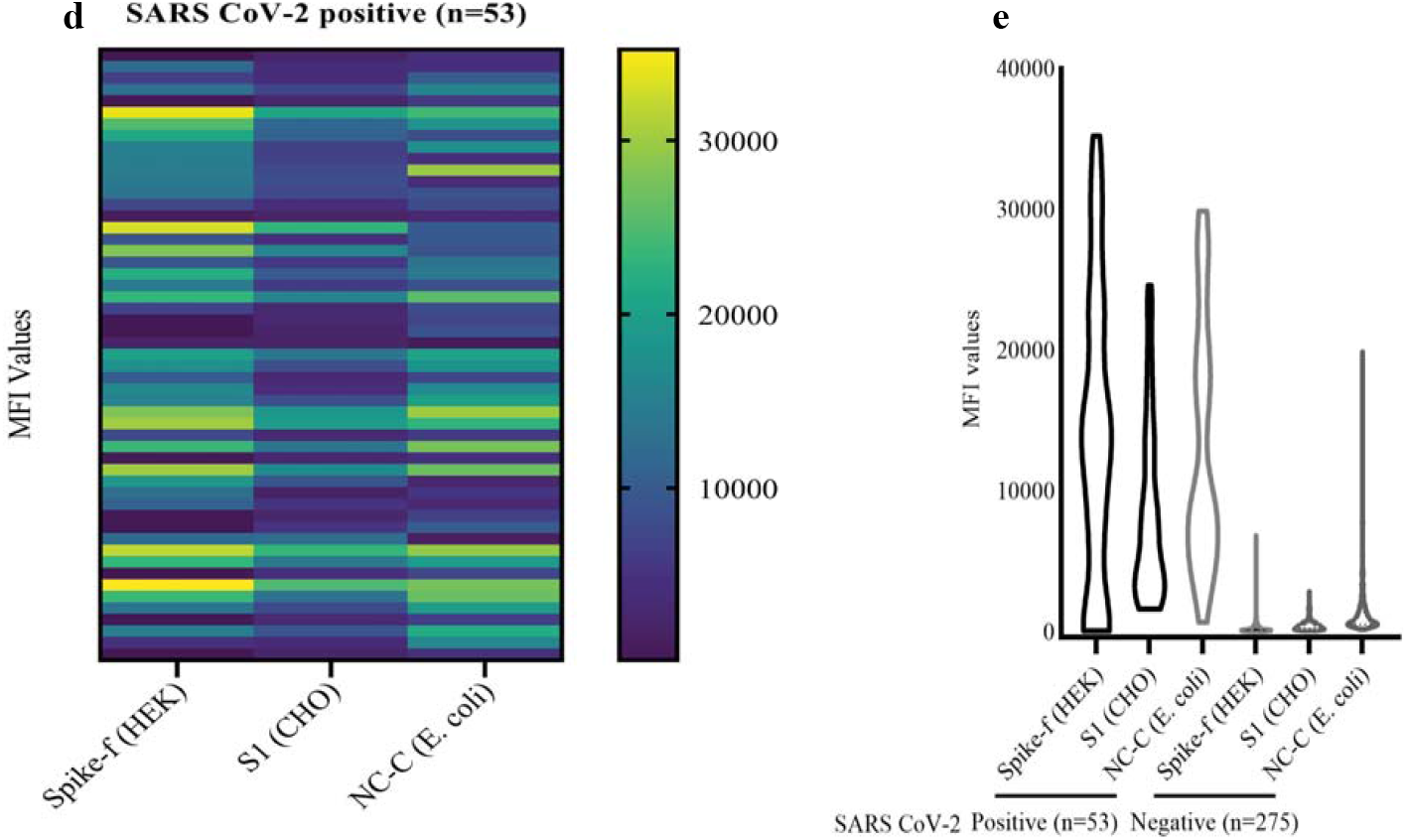
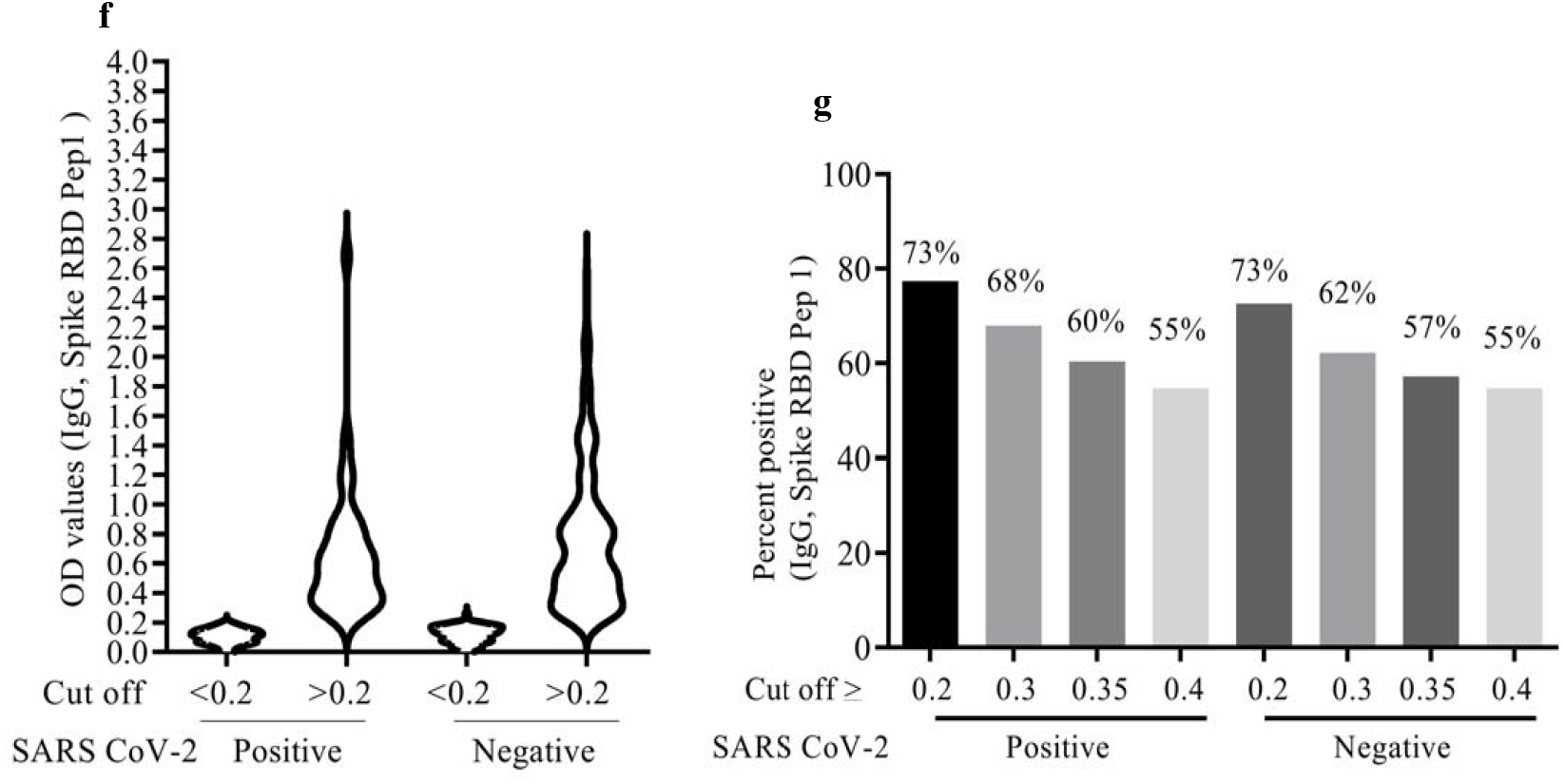
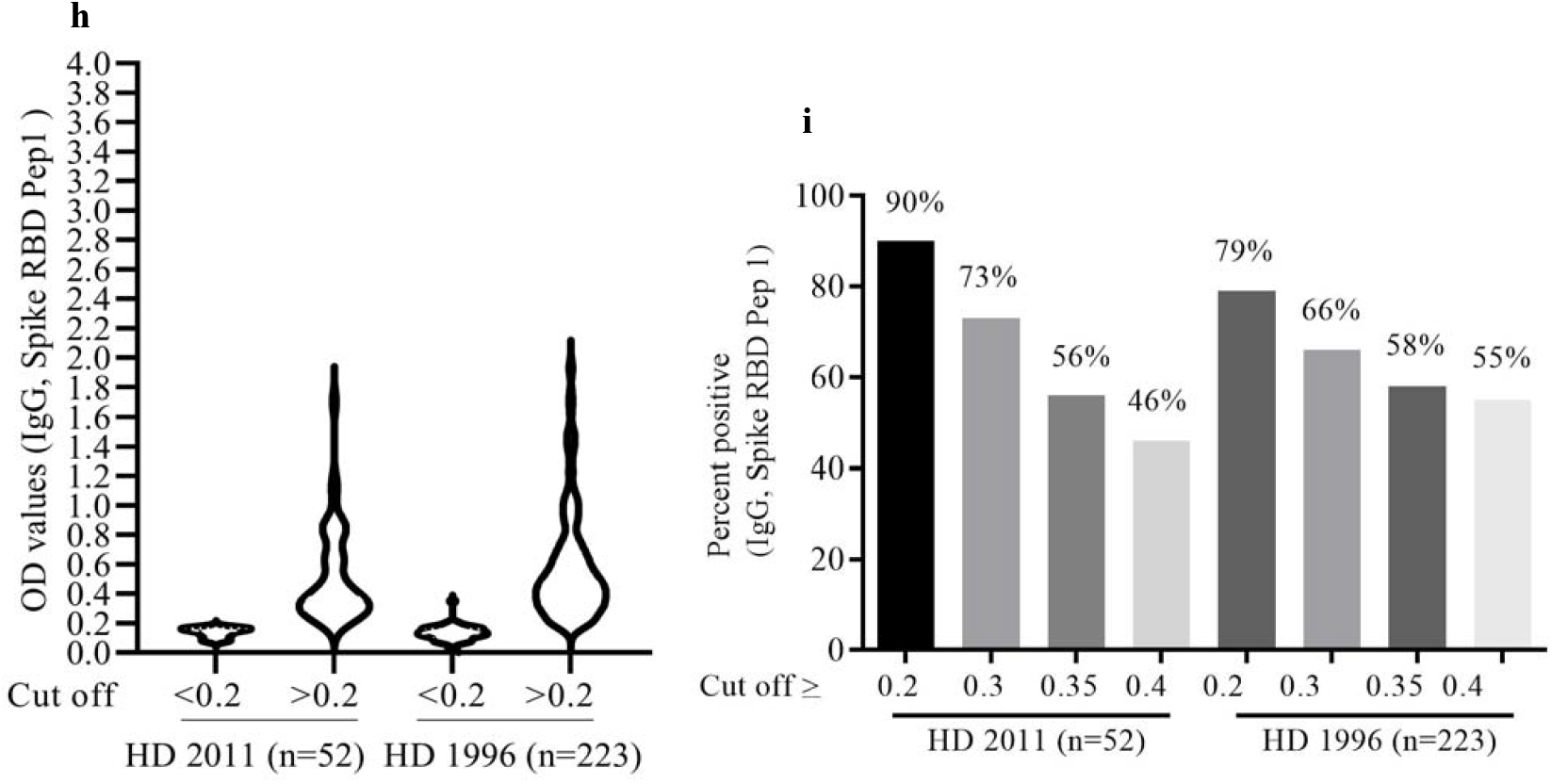
Localization of peptide sequences within spike protein of SARS-CoV-2. (a) Localization of the NGVEGF peptide in the spike protein of SARS-CoV-2. (b) NGVEGF is present in the critical domain of SARS-CoV-2 that interacts with the ACE2 receptor. (c) NGVKGF is situated in a highly immunodominant region of the neuraminidase enzyme of influenza A H1N1 and is expected to elicit an antibody response in most people infected with influenza A H1N1. **Receptor binding motif–specific antibodies are present in COVID-19-negative unvaccinated individuals**. (d and e) Fifty-three of 328 subjects had IgG antibodies (mean MFI >6 SD) against soluble pre-fusion stabilized trimeric spike glycoprotein (SPIKE-f (HEK). (f) NGVEGF-specific IgG antibody levels in COVID-19-positive and COVID-19-negative subjects, measured by ELISA. (g) Prevalence of NGVEGF-specific IgG antibody values (cut-off values are shown for OD >0.2, >0.3, >0.35, and >0.4) in COVID-19-positive and COVID-19-negative subjects. (h) NGVEGF-specific IgG antibody levels in serum samples from healthy donors in 2001 and 1996, measured by ELISA. (i) Prevalence of NGVEGF-specific IgG antibody values (cut-off values are shown for OD >0.2, >0.3, >0.35, and >0.4) in healthy donors (HD) from 2011 and 1996. OD: optical density,

Remarkably and very interesting, the NGVEGF/NGVKGF peptide was present in the most critical part of the receptor binding motif of the spike protein (amino acids (aa) N481 to F486, Fig. 1a) that interacts with the angiotensin converting enzyme 2 (ACE2) receptor (Fig. 1b) (37). In H1N1, the NGVEGF/NGVKGF peptide is situated in a known immunodominant region of the neuraminidase protein (Fig. 1c) and is hence expected to elicit an antibody response in a high proportion of people infected with influenza A H1N1 strains after 2008 (38). Flu strains containing NGVKGF are currently circulating around the globe and have been included in seasonal flu vaccines during the last decade. It is therefore possible that antibodies to NGVEGF/NGVKGF developed during an Influenza A H1N1 infection or after a seasonal flu vaccination could have protected some people against pre-Omicron strains of SARS-CoV-2.

### Receptor binding motif–specific antibodies are present in COVID-19-negative unvaccinated individuals

To determine if NGVEGF peptide specific antibodies are present in healthy unvaccinated COVID-19 negative individuals, we collected plasma/serum samples from 328 healthy persons in September 2020 and analyzed the samples for the presence of IgG-specific antibodies to SARS-CoV-2 spike and to the NGVEGF peptide, and developed a diagnostic ELISA method to detect NGVEGF peptide– specific antibodies. Spike-specific antibodies were detected in 53 (16.2%) of 328 samples (Fig. 1d and e) (30) with a multiplex assay and determined who among the donors that were positive or negative for SARS-CoV-2.

Since most people have been exposed to influenza A H1N1 strains since 2008, and flu specific antibodies can be passively transferred to infants via placenta before birth, we had no optimal negative control samples to estimate a strict cut-off value for positivity in the ELISA test. Instead, we relied on negative controls for peptide reactivity of irrelevant peptides, and also confirmed a lack of NGVEGF specific antibodies in mice plasma (S Fig. 1). In mice, flu vaccination elicited development of NGVEGF peptide specific IgM and IgG antibodies (S Fig. 1 a and b); these mice serum samples also served as positive and negative controls for the ELISA test. For the human sera, the prevalence of IgG positivity to NGVEGF was 73% at a threshold optical density (OD) value ≥0.2 (Fig. 1F and g), 68% at OD ≥0.3, and 55% at OD ≥0.4. Of note, only one of 53 COVID-19-positive subjects had high titers of NGVEGF-specific antibodies (OD >2, Fig. 1f).

We next established a Luminex Multiplex bead array assay to compare antibody reactivity to 8 NGVEGF or NGVKGF spike peptides (11 or 17 aa in length, Table S2). The peptides were synthesized with a cysteine bridge and biotinylated at the N or C terminus, respectively. Three SARS-CoV-2 peptides, an adenovirus peptide, and an irrelevant peptide (Neglle1) served as controls. Using this method, we confirmed a variable antibody reactivity to NGVEGF (peptide 3) and NGVKGF (peptide 7) in human sera from different individuals (Fig. 2a–d). The highest reactivity was observed to the longer NGVKGF peptide biotinylated at the N terminus containing the cysteine bridge (peptide 7) and was found both in COVID-19-positive and -negative individuals (Fig. 2b and d). Sera that contained NGVKGF- or NGVEGF-reactive antibodies however rarely recognized the full-length recombinant spike protein. COVID-19-positive subjects generally had low median absolute deviation (MAD) values to NGVKGF (peptide 7, Fig. 2 e and f), and some did not mount an antibody response to NGVEGF (peptide 3, Fig. 2e). However, several subjects with anamnestic flu and some with known family exposure to SARS-CoV-2 who did not become infected with SARS-CoV-2 in spite of exposure, interestingly had antibodies with high MAD values to NGVKGF (example in Fig. 2g). High levels of NGVKGF and NGVEGF specific antibodies were also prevalent among COVID-19-negative blood donors with unknown SARS-CoV-2 exposure (example in Fig. 2h).

**Figure. 2.**
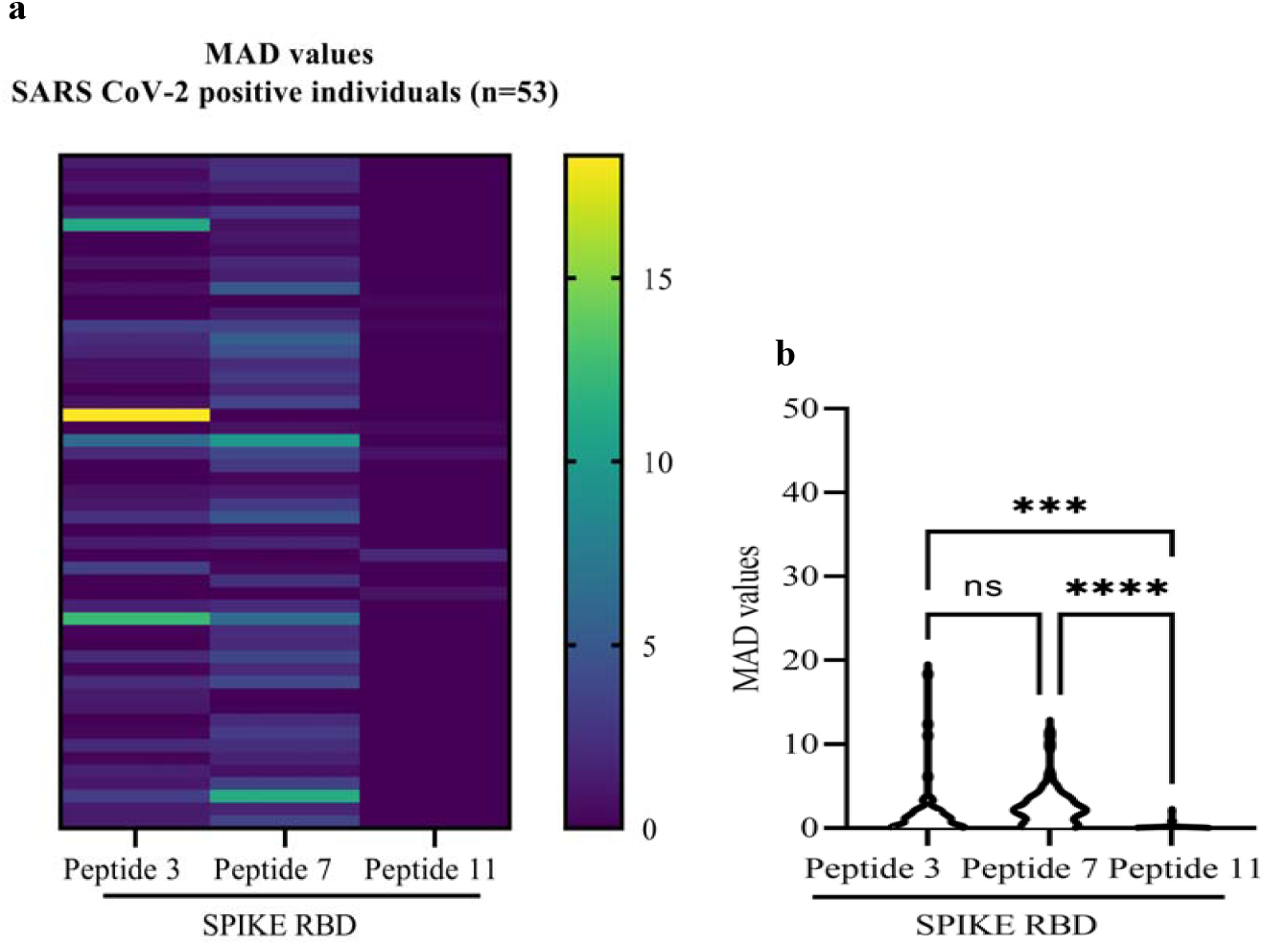
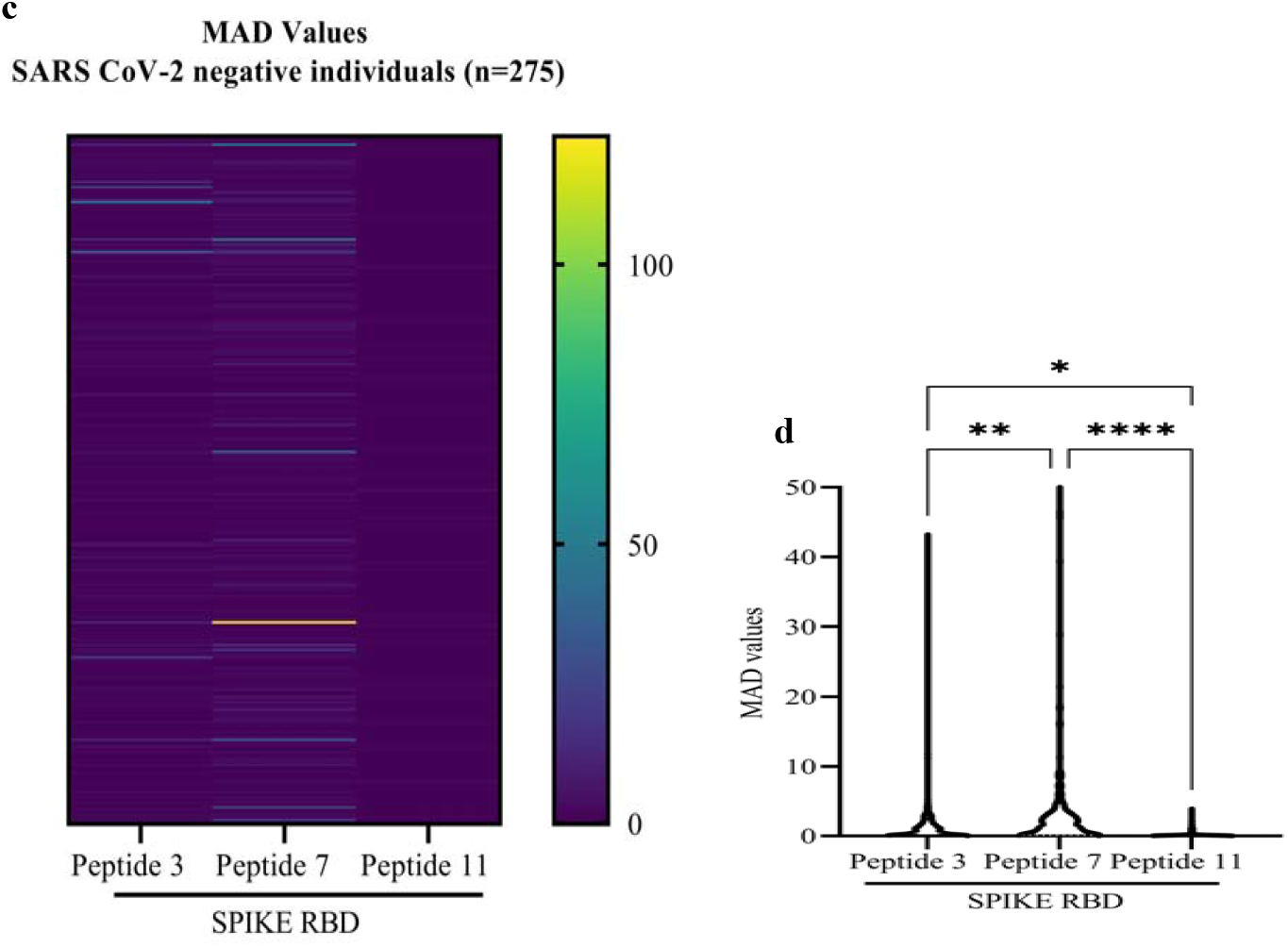
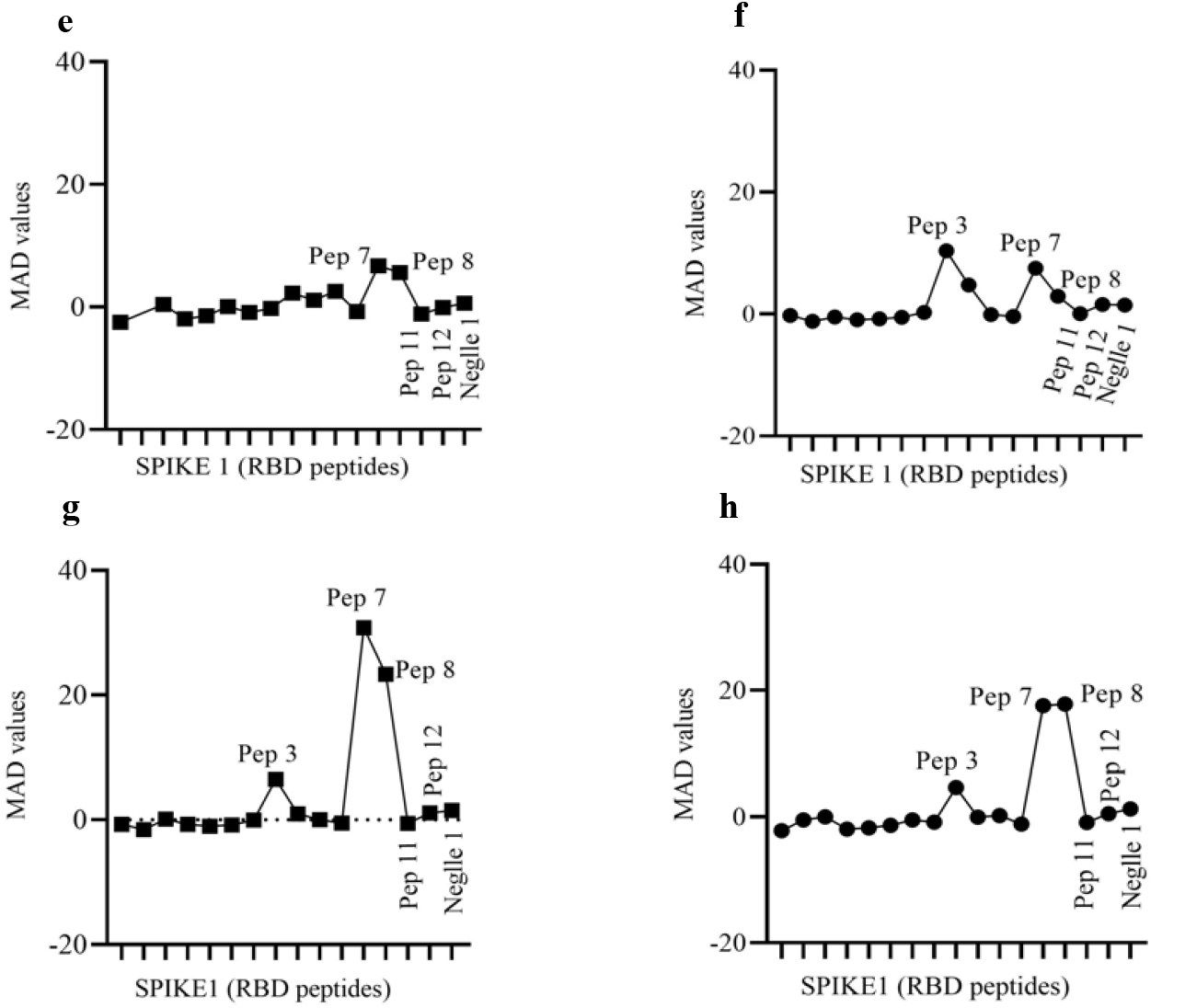
NGVEGF- and NGVKGF-specific IgG antibodies are present in both COVID-19-positive and COVID-19-negative individuals. (a and c) Heat map of MAD values for peptides 3, 7, and 11 in COVID-19 positive (A) and COVID-19-negative (c) cohorts. (b and d) The IgG antibody levels (MAD values) to the NGVKGF (peptide 7) were higher than to the NGVEGF peptide (peptide 3) in both COVID-19-positive (b) and COVID-19-negative (d) cohorts. (e–h) Peptide-specific IgG antibodies in two COVID-19-positive individuals (e and f), in a subject who was exposed to Flu and SARS CoV-2 but did not get sick (g), and in an COVID-19-negative subject (h). One-way ANOVA and Tukey’s multiple comparison test were used for multiple comparisons. ****p<0.00001, ***p<0.0001, **p<0.001, *p<0.01, ns: no significant.

The NGVEGF peptide arose in swine flu in 2009 and the variant NGVKGF was subsequently present in >99% of H1N1 strains. The NGVKGF peptide was also present in 31.4% of strains (*n* = 1467) sequenced before 2008. We hypothesized that these peptides could have generated cross-protective immunity to SARS-CoV-2, and therefore examined the prevalence of NGVEGF-reactive antibodies in patient sera collected before and after 2008 (52 samples from 2011 and 223 from 1996). At an OD ≥0.2, 90% of sera from 2011 and 79% of sera from 1996 (Fig. 1h and i) contained NGVEGF-reactive IgG antibodies (Fig. 1i). With a cut off of OD ≥0.3, 73% of sera from 2011 and 66% of sera from 1996 (Fig. 1h and i) contained NGVEGF-reactive IgG antibodies. Since antibody prevalence to NGVEGF was higher than expected in serum from 1996, we searched for other peptides that could have elicited an antibody response to the NGVEGF peptide that could cross-react with the receptor binding domain of SARS-CoV-2. By structural analyses, we identified two additional peptides—DGVKGF, and NGIKGF—that were similar to NGVEGF and hence could also have contributed to a protective immune response to SARS-CoV-2. NGIKGF was only present in 1 (<0.01%) and DGVKGF in 990 (67.5%) of 1467 H1N1 strains before 2008 (Table S1). After 2008, DGVKGF was present in 107 (0.56%) and NGIKGF in 12 (<0.01%) of 18,972 sequence H1N1 strains. Thus, the DGVKGF, NGVEGF, and NGVKGF peptides have been present in many influenza A H1N1 strains over long periods of time and these peptides may have triggered antibody responses mediating specific cross-protective immunity to the receptor binding motif of the SARS-CoV-2 Spike protein.

### Inhibitory antibodies to SARS-CoV-2 and NGVEGF specific T cells can increase after flu vaccination

To examine if NGVEGF specific antibody titers and NGVEGF specific T cells increase in flu and COVID-19 vaccinated individuals and if these could neutralize SARS-CoV-2 infection, we collected plasma and blood cells from 20 individuals before and after Flu and after COVID-19 vaccination. Plasma samples from only 19 subjects were available for this test, as when ethical permission was granted for the study in December of 2020, almost all flu vaccines had been administered in Sweden, and COVID-19 vaccinations were ready to start. We first tested plasma samples from these individuals collected before and after flu and COVID vaccination for the presence of NGVEGF specific antibodies. We observed a trend for enhanced IgG titers in 7 healthy individuals, but not among 12 elderly people who lived in an elderly care home and were considered “vulnerable due to poor health” (Fig 3a and b). In all subjects, the NGVEGF specific antibodies decreased after COVID-19 vaccination. We next tested the neutralizing capacity of antibodies in these plasmas in a virus neutralization cell culture test, but the plasma samples were toxic to the cultured cells and resulted in high cell death. This phenomenon was not observed when control sera were tested in the same assay. As the plasmas were not possible to use in this cell culture assay aiming to test if antibodies present in the plasma had an inhibitory effect on SARS-CoV-2 infection, we examined the ability of plasma containing NGVEGF/NGVKGF specific antibodies to inhibit binding of the spike protein to the ACE2 receptor in a SARS-CoV-2 surrogate virus neutralization test (sVNT). We observed some enhanced potential protective SARS-CoV-2 immunity by flu vaccination (VaxigripTetra Quadrivalent Flu vaccine, Sanofi Pasteur (39) as measured by enhanced NGVEGF specific antibodies and the SARS-CoV-2 sVNT (Figure 3 c, d, e, and Table S5). Although the data obtained from these analyses are limited and should only be considered descriptive in its nature, we made some interesting observations. Three subjects with no recent anamnestic flu infection had low binding inhibitory activity (mean 32.7% inhibition) that was enhanced to mean 55% by flu vaccination) and this inhibitory capacity was further increased by COVID-19 vaccination (to mean 94%) (Fig. 3c), even though the NGVEGF specific antibodies were decreased (Figure 3a). Four subjects with suspected flu infection within the past 2 years, had inhibitory antibody activity at levels similar to those after flu vaccination (mean 47% neutralization), and this neutralizing capacity did not increase more by flu vaccination (mean 47% inhibition), but was further enhanced by COVID-19 vaccination (to mean 72%) (Fig. 3d). Twelve of the subjects lived in an elderly care home and were considered “vulnerable due to poor health” and only had a minor increase in the inhibitory effect of antibodies after flu vaccination, from mean 34% before to a mean of 40% after flu vaccination and to 61% after COVID-19 vaccination (Fig. 3e and Table S5); for some of them the neutralizing capacity to SARS-CoV2 spike was still concerningly low (Fig. 3e and Table S5). Nine (75%) had an adequate response to the mRNA BNT162b2 COVID-19 vaccine from Pfizer-BioNTech (mean inhibition 68%), and three had an insufficient response (mean 39% inhibition) after two vaccine doses (Fig. 3f and Table S5). This was also reflected in lower SARS-CoV-2 specific antibody levels. This inhibitory effect was higher in the seven other subjects of various ages, and increased from 41% before to 51% after flu vaccination and to 81% after COVID-19 vaccination (n=7, Fig. 3g), and they all had adequate antibody levels to SARS-CoV-2 after vaccination.

**Figure. 3.**
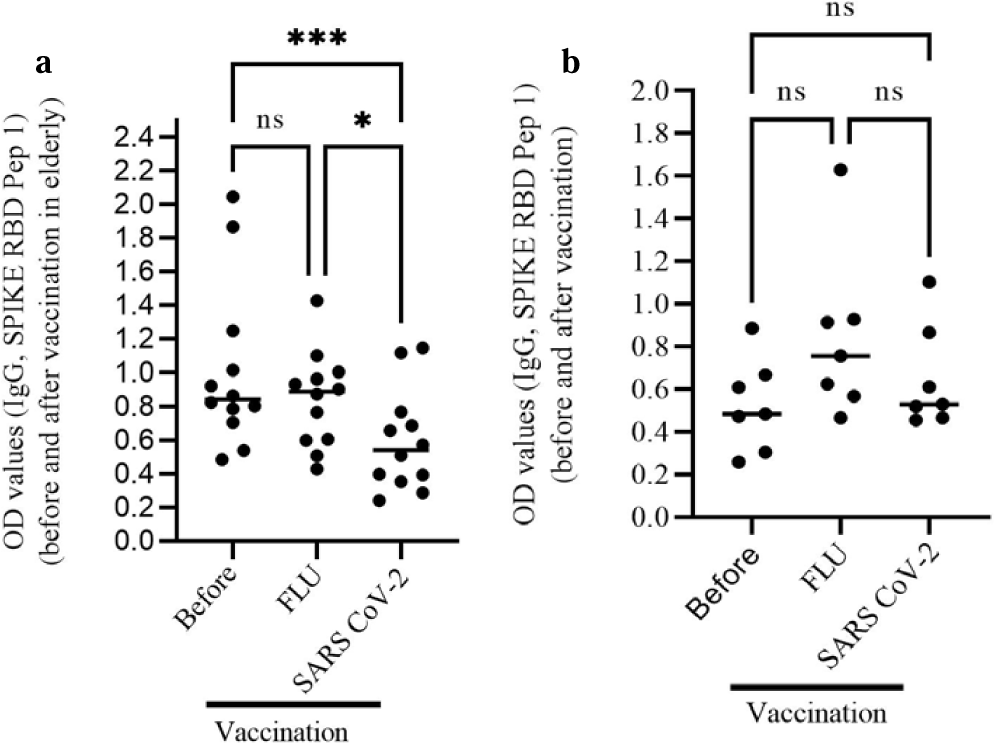
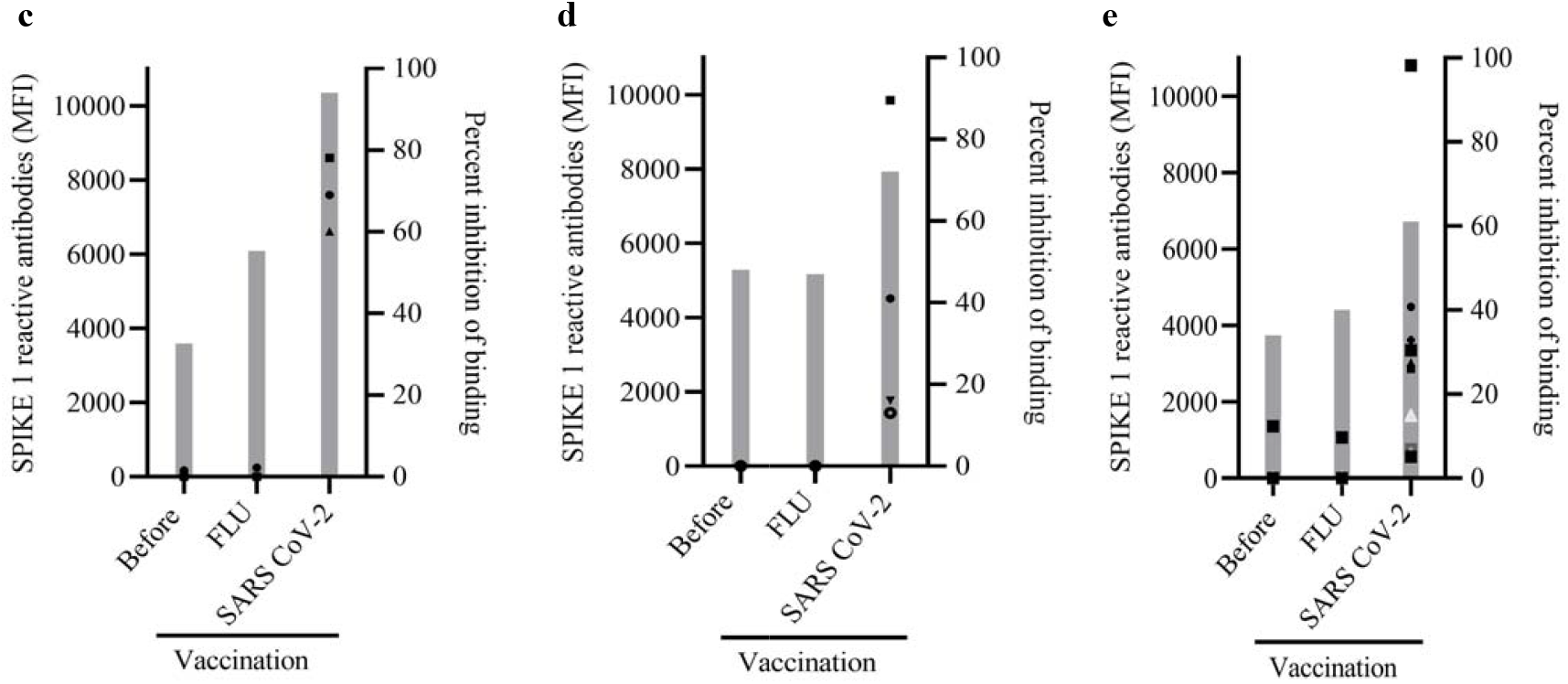
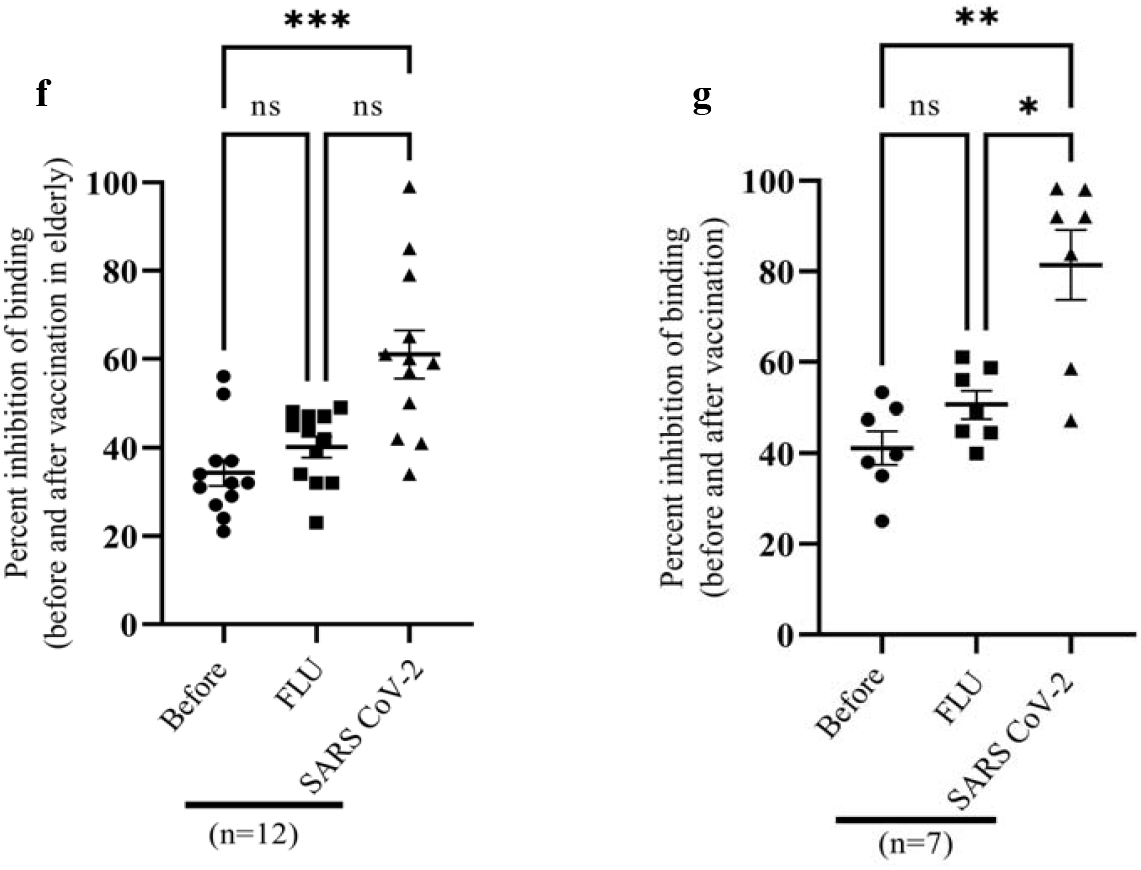
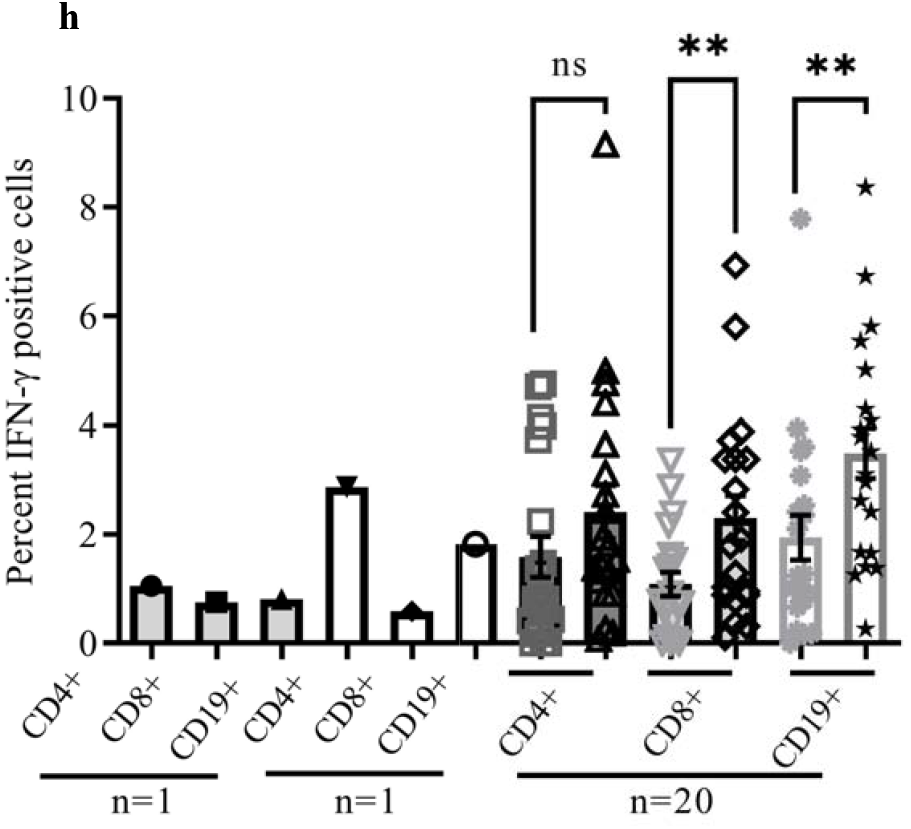
Inhibitory antibodies to SARS-CoV-2 and NGVEGF specific T cells can increase after flu vaccination. (a and b) Plasma from 19 individuals before and after Flu and after COVID-19 vaccination were examined for presence of NGVEGF specific antibodies using ELISA. A trend was observed for enhanced IgG titers in 7 healthy individuals, but not among 12 elderly people who lived in an elderly care home and were considered “vulnerable due to poor health”. (a and b) In all subjects, the NGVEGF specific antibodies decreased after COVID-19 vaccination. (c-e) Inhibitory binding of Spike to the ACE2 receptor detected in plasma/serum with a surrogate virus neutralization assay: Grey bars represent mean value of binding inhibition; individual signs represent MFI value for Spike 1 reactive antibodies. (c) Individuals (*n* = 3) with no recent anamnestic flu infection had lower binding inhibitory activity, which was boosted by flu vaccination. (d) Four subjects who had evidence of anamnestic flu had binding inhibitory activity at levels similar to those after flu vaccination; this immunity level was not further enhanced by flu vaccination. (e) In 12 subjects, COVID-19 vaccination increased binding inhibitory activity against SARS-CoV-2 to a mean value of 61%. (f) Increased binding inhibitory activity after COVID-19 vaccination (mean values: 34.3% before vaccination, 40.2% after flu vaccination, and 61% after SARS CoV-2 vaccination) in elderly cohort in care home (g) and in healthy donors (mean values: 41.2% before vaccination, 50.6% after flu vaccination, and 81.4% after COVID-19 vaccination). Expansion of IFN-γ producing NGVEGF peptide specific T cells in Flu-vaccinated subjects. (h) Increased number of IFN-γ-producing B and T cells stimulated with NGVEGF peptides and analyzed by flow cytometry in a group of 20 subjects before and after seasonal flu vaccination (CD4, CD8, and CD19 cells). Nonparametric One-way ANOVA test and Dunn`s multiple comparison test were used for multiple comparisons of percent of antibody and antibody-binding inhibition before and after vaccinations. Paired *student t-test* was used for comparison of IFN-γ producing cells before and after Flu vaccination.

### Flu-mediated protection to SARS-CoV-2 may vary in different populations

While antibodies may protect people from becoming infected, cytotoxic T cells are crucial to resolve life-threatening infections by killing virus-infected cells. In modeling analyses, we found that the NGVEGF peptide in theory can be presented to CD8 T cells by some HLA class I molecules (HLA-A*33:01, HLA-A*68:01, HLA-A*31:01, HLA-A*11:01); these are found in about 22.2% of Scandinavians (Table S3). We confirmed that B and T cells from 20 healthy subjects recognized and responded to NGVEGF peptides *in vitro* and that this reactivity was boosted in some people after seasonal flu vaccination (Fig. 3h). This response was highly individual and differed for CD19, CD4, and CD8 T cells. Flu-vaccinated subjects had significantly more IFN-γ producing CD8 T cells that recognized the NGVEGF peptide (mean increase from 1.1% to 2.3%, *p* = 0.009) and B cells (mean increase from 1.9% to 3.5%, *p* = 0.003); IFN-γ producing CD4 T cells reactive to the NGVEGF peptide also trended higher after flu vaccination (mean increase from 1.6% to 2.0%, *p* = 0.0567, Fig. 3h). Interestingly, 7 of 20 (35%) individuals had a robust increase in the numbers of CD8 T cells reactive to the NGVEGF peptide (mean increase of 4.3%); we predicted that about 22% of Scandinavians have HLA types able to present this peptide efficiently to T cells. Thus, the number of T cells reactive against the NGVEGF peptide increased prominently in some individuals after Flu vaccination.

Further peptide screening identified 11 additional influenza H1N1 cross-reactive CD8 T-cell peptides to SARS-CoV-2 (Table S4). Modeling implied that they could be presented by HLA types found in about 71% of people in Scandinavia (mainly HLA-A*02:01 and HLA-A*01:01), but in only 40% of people worldwide (Table S3). These observations suggest that the strength of protective immunity induced by influenza A H1N1 strains that could have mediated protection against SARS-CoV-2 may vary around the globe.

### Mathematical modeling supports that the existence of a pre-existing immunity to SARS-CoV-2 dampened pandemic spread of the virus before Omicron emerged

Evidently, a substantial proportion of the world’s population had antibodies to NGVEGF and T cells reactive against flu peptides that could have provided protection from infection or severe COVID-19 disease during the first waves of the pandemic, and before vaccinations were introduced. To further understand whether a pre-existing flu-mediated cross-immunity to SARS-CoV-2 could had dampened the epidemic on a population level, we turned to mathematical models.

We implemented the SEIR-code by Britton et al. (40), of which there are two versions. The first version takes interactive patterns between different age groups into account (Age SEIR), and the second also considers variations in social activity (Age-Act SEIR) (Fig. 4a). We tested these models on case data from Stockholm, using data from the second wave (Fig. 4a and b), as testing was not reliable during the first wave. Data from the Swedish Public Health Agency allowed for separation of cases by the original versus the alpha strain. The blue curve in Fig. 4 represents cases in the second wave caused by the original strain only, readjusted to account for underreporting of cases. Based on data from the Swedish Public Health Agency, seroprevalence was 10% in Stockholm at the start of the second wave (early September 2020) and rose to 22.6% in mid-February 2021, between the second and third waves (11, 12). These findings are consistent with the seroprevalence estimated from our serology data: 16.2% (*n* = 328) in late September 2020 and 21.1% (*n* = 450) in late February 2021. When we attempted to fit the curve of cases with either SEIR model using an immunity level of 10%, the curves are nowhere near reality (Fig. 4a). However, an almost perfect fit to measured data was observed when we used pre-immunity levels of 60% for Age-SEIR and 50% for Age-Act SEIR, which corresponds to a pre-pandemic immunity protective level of 60–70% (Fig. 4b). This level correlates remarkably well with the measured seroprevalence of NGVEGF-specific antibodies in people in Stockholm (55–73%, depending on OD cut-off for positives) and with the modeled HLA class I–mediated protective T-cell immunity levels for a Scandinavian population (estimated to be 71% according to expected HLA types in the population). No other parameter we examined affected the model output in a similar manner. Thus, it was not possible to match modeled data to actual case data without taking a substantial protective pre-existing immunity into consideration (Fig. 4a and b), and the modelled data corresponded remarkably well with the measured pre-immunity levels to the Influenza A H1N1.

**Figure. 4.**
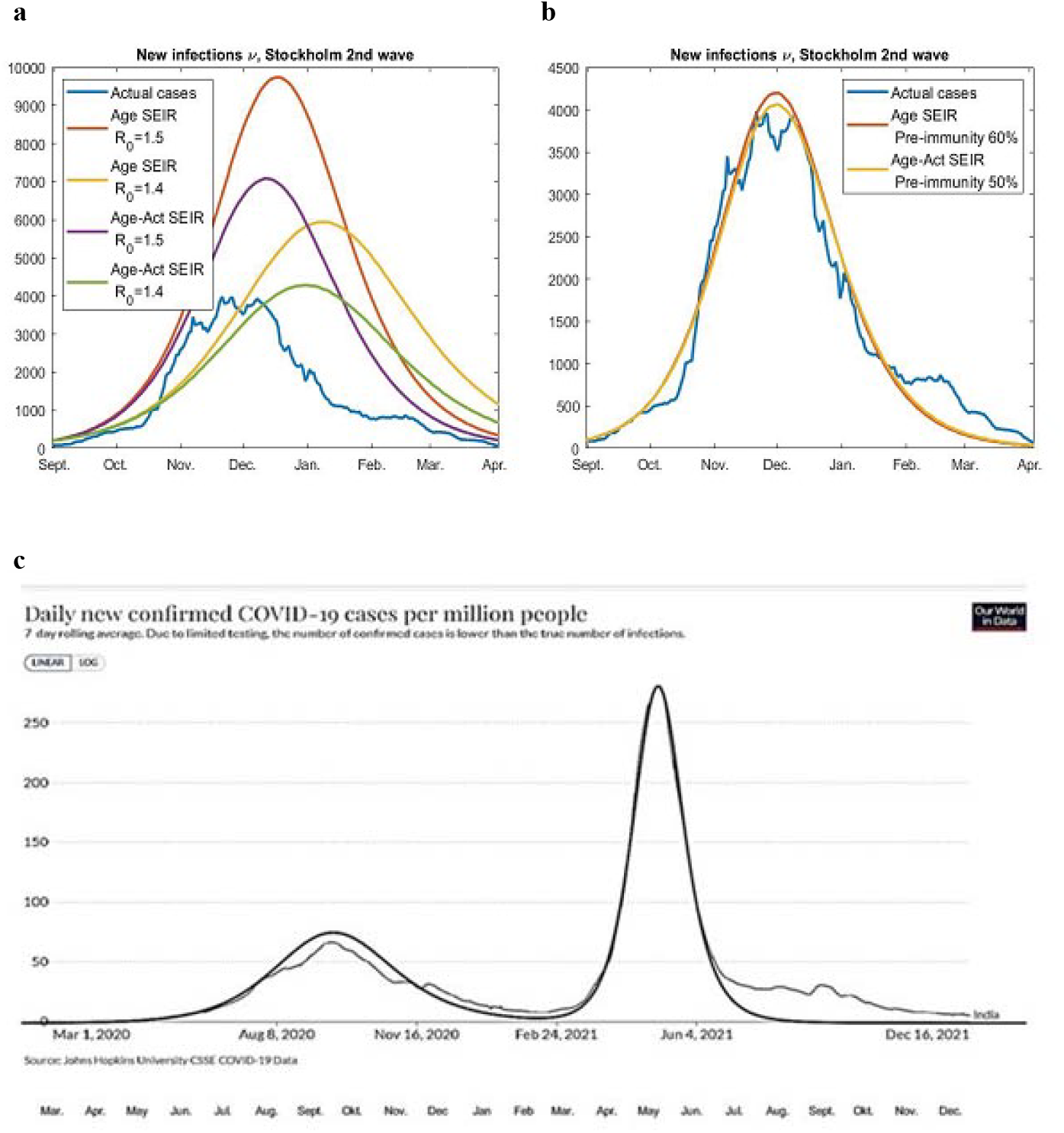
Mathematical modeling considering pre-existing immunity predicts COVID-19 outbreaks. Without taking pre-immunity into account, it was not possible to match the development of the second wave in Stockholm County with two heterogeneous SEIR-models developed by Britton et. al.(40): the Age-SEIR model, which takes variable social interactions between different age groups into account, and the Age-Act-SEIR, which also takes variations in social activity within each age group into account. Blue curves are for actual cases. (a) Attempts to fit actual cases in the absence of pre-immunity, using different R_0_ values. (b) Curves generated with Age-SEIR using 60% pre-immunity and with Age-Act-SEIR using 50% pre-immunity. (c) The pandemic progression can be accurately modeled for India, using a pre-immunity of 25% against the Wuhan strain and 10% against the Delta variant.

We next ran the code on case data for India, which we expected had a lower pre-immunity protection, as a result of their populations different HLA types compared to Scandinavians. A pre-immunity protection in India is therefore expected to be effective at around 20–30% in the first Wuhan strain wave and only be protective at a 10–20% level for India’s delta wave). We used seroprevalence data released by the Indian Council of Medical Research; 7% in August/September 2020, 24% around January 1, 2021, and 67% in June/July 2021 (https://indianexpress.com/article/explained/explained-icmr-covid-fourth-serosurvey-findings-7413949/). Using a 25% pre-immunity against the original strain and a 10% pre-immunity for the delta variant, and assuming an antibody half-life of 16 months after natural infection (41), the model fit the observed case data very well, implying that pre-immunity protection, as we hypothesized, was lower for India than Stockholm. These observations give further validation to the real-life value of our findings, i.e that a protection to SARS-CoV-2 could have been mediated by previous Influenza A H1N1 infections and that this protection may be different in different populations who have different HLA types and thereby variable capacity to present flu peptides to T cells.

## Discussion

We discovered that the receptor-biding motif in the SARS-CoV-2 spike protein that interacts with the ACE2 receptor is identical or very similar in structure to highly immunogenic peptides, NGVEGF, NGVKGF and DGVKGF that are present in neuraminidase of many influenza A H1N1 strains that have circulated around the globe during the last decades. We provide evidence that 55%–73% of individuals have pre-pandemic existing antibodies to this peptide that could have mediated some protection against SARS-CoV-2, and we provide evidence that flu vaccination can increase immune protection to NGVEGF and hence potentially against SARS-CoV-2. Since a majority of people have some NGVEGF-reactive antibodies, these new insights also affect the interpretation of the role of NGVEGF-specific antibodies in SARS CoV-2 infected individuals, especially concerning their protective effects against variant viral strains containing the E484K, E484Q or the new E484A omicron mutation (42). Our modeling data give further support to the hypothesis that pre-existing immunity to influenza A H1N1 strains indeed could have protected a large set of people from SARS-CoV-2 infection or severe COVID-19 before omicron emerged in Scandinavia, whereas this protection may have been less prominent in India with a population carrying other HLA types. Indeed, India had a much worse second wave than Sweden. Importantly, this discovery provides explanations to the epidemiological observations that seasonal flu vaccination appears to provide significant protection against COVID-19 infections, hospitalizations, ICU admission, and death (43-48); with estimated protection rates (40-80%) that are in range of what was estimated in the present study (55-73%).

The identified cross-reactive pre-immunity is not expected to provide sterilizing immunity to SARS-CoV-2. Rather, we mainly consider that it acted as a brake on the epidemic viral spread, as a higher viral dose was needed to infect someone who had a substantial level of flu-mediated pre-immunity under given non-pharmacological interventions. However, we also consider the possibility that those individuals could have been better protected from severe COVID-19 disease. People with high titers of antibodies and higher levels of T cells that cross-react with flu and SARS-CoV-2 were likely better protected against SARS-CoV-2 before Omicron emerged, perhaps especially if they had a recent H1N1 infection or seasonal flu vaccination. This hypothesis is supported by the observed increased specific immune activity to NGVEGF in some flu vaccinated individuals in the present study. Whether a person who had flu-mediated protective immunity became infected or not, and whether such individual developed severe diseases or not may have depended on the infectious dose and the level of immunological protection against flu at the time of exposure.

On a population level, this cross-protective immunity may explain the unexpectedly slow unfolding of the pandemic in Sweden despite the absence of a lock-down, while India that we predicted had a lower efficiency of the flu-mediated protection, was hit harder by the early SARS-CoV-2 strains. The relevance and accuracy of this theory was strengthened by our modelling data. One can then speculate over the following scenario; if this cross-protective immunity did not exist, could we have experienced as rapid spread of the original strain as Omicron, which appears to avoid both natural immunity, vaccination immunity and influenza pre-immunity. If the spread of the more pathogenic Wuhan strain causing more severe disease would have been as fast as omicron, the pandemic could have been even more devastating than so far observed.

Mutant strains have rapidly outcompeted the original strain and new “variants of concern” that are more contagious have spread rapidly over the world, as natural immunity, vaccine immunity and potentially flu mediate pre-immunity provide less protection against them (49, 50). Is it then possible that flu-mediated immunity has selected for the early variants of concern? The Gamma (P1, Brazil), Alpha (B1.1.7, Britain), and Beta (B.1.351, South Africa) strains contain an N501Y mutation that is thought to enhance by 10-fold the binding affinity of the spike 1 protein for the ACE2 receptor (51-53). The Delta strain (B.1.617) has a T478K mutation, and the NY Iota strain has a S477N mutation in the receptor binding domain, which was predicted to reduce protection from vaccines (54, 55). Interestingly, the N501Y, S477N, and T478K mutations flank the NGVEGF cysteine loop at aa 481 to 486, which interacts with the ACE2 receptor. As a result of these mutations, NGVKGF-specific antibodies may have become less protective, enabling these viral strains to infect a higher proportion of individuals at lower doses. It is then possible that the N501Y, S477N, and T487K mutations in SARS-CoV-2 evolved through laws of Darwinian evolution to increase affinity for the ACE2 receptor and to evade NGVEGF-interacting antibodies, making these variants more contagious? Under such scenario, the pre-existing flu-mediated immunity may have aided in the selection of the first new viral variants of concern.

Our data also opens for speculations of the relevance of this finding for SARS-CoV-2 susceptibility especially among children. High numbers of hospitalized children infected with the delta variant were reported in the US, the UK, and Israel, but not in Sweden. One could speculate over the possibility that flu vaccine strategies could have affected SARS-CoV-2 severity among children. Seasonal flu vaccinations are recommended for children in the US, the UK, and Israel, but not in Sweden. In Sweden, flu vaccinations are only recommended to high-risk groups and for people over 65 years of age. A flu-mediated pre-immunity to SARS-CoV-2 may be less efficient and less sustainable after repeated flu vaccinations than after influenza A H1N1 infection that would elicit a more robust B- and T-cell immunity. The flu vaccination recommendations and favorable HLA types might explain Sweden’s lower incidence of severe COVID-19 disease in children under 10 years of age of which many had swine flu as their first influenza A H1N1 infection, and hence developed a robust immune response to NGVEGF/NGVKGF peptides. Before Omicron, Sweden did not have high rates of severe infections among children requiring hospital care; however, multisystem inflammatory syndrome in children was twice as prevalent in Sweden as in the US in November 2021. This syndrome develops 4–6 weeks after diagnosis of COVID-19 and does not seem to be related to the severity of SARS-CoV-2 infection; its incidence may rather be related to the number of infections. Sweden kept schools open during the pandemic and had high levels of transmission among children, but few became severely ill in the acute phase of COVID-19. Flu vaccination strategies together with unfavorable HLA types with lower capacity to present flu peptides to T cells may hence explain differences in susceptibility to SARS-CoV-2 infection and the risk of developing severe COVID-19 in different parts of the world. We observed that many children below 10 years of age were hospitalized in Sweden during the Omicron wave, which was not observed during earlier wave. This may be explained by their loss in previous influenza mediated protection due to the E484A Omicron mutation.

Our experimental and modeling data have limitations. First, levels of antibodies against NGVEGF, and the neutralization assay have no threshold to estimate protection on an individual level and antibody protection should be considered to be more relevant on a population level, and we had no suitable NGVEGF antibody negative human sera to use as a control. We have searched for control sera and T cells from 6-9 months old children, as these should be predicted to have the lowest prevalence of NGVEGV specific antibodies and T cell reactivities, but found no such samples among contacted colleagues. Second, the mathematical model can only suggest a range of pre-immunity levels that are likely to be true in reality. Both SEIR models we used in earlier work (11, 12) and the method developed here are crude tools, and the results should be interpreted with caution. However, SIR (susceptible, infective, recovered) and SEIR models that did not include pre-immunity completely failed to predict the dynamics of SARS-CoV-2 spread (11, 12); pre-immunity levels that we here found to be different in Stockholm versus in India. This is the main mathematical argument for the existence of a pre-existing immunity, the exact level of which is hard to estimate with certainty. On the other hand, the estimated pre-immunity levels from SEIR models and the completely different mathematical tool we devised for this study yielded remarkably consistent results, near the levels suggested by pre-immunity data observed here—supporting the existence of flu-mediated antibodies and T cells that cross react with and protect against SARS-CoV-2 infection or from severe COVID-19 disease in a high proportion of Swedish people. This scenario would also explain why so many people in Sweden were not infected despite household exposure, had asymptomatic infections, or experienced mild disease before Omicron emerged.

We conclude that the high prevalence of flu-mediated cross-protective immunity to SARS-CoV-2 is important for understanding SARS-CoV-2 susceptibility, vaccine responses, protection against new variants, the natural course of COVID-19 in different individuals, as well as for the impact of this virus and its mutants on people and our society. Learnings from such pre-immunity protection studies are important to consider to better handle future pandemics.

## Data Availability

All data produced in the present study are available upon reasonable request to the authors

## Ethics Statement

The study was approved by the Ethics Committee at Karolinska Institutet, Stockholm, Sweden (Dnr 2020-06333: all vaccinated subjects gave written informed consent; blood donors from blood bank were anonymous; Dnr 2020-07232: all subjects gave written informed consent; Dnr 06400: included anonymous blood samples from healthy donors (HD) in 2011 (Dnr: 01-420) and in 1996 (Dnr: 95-397 and 02-091). The study protocol for animal study was approved by II Local Ethics Committee for Animal Experiments in Warsaw (Dnr: 16.07.2021).

## Author Contributions

Conceptualization: CSN, MC, AS, BS. Methodology: CSN, AR, BR, PR, AS, SA, ÅL, TH, MO, BS, CM, MC, LK, NMA. Software: MC. Visualization: AR, PR, AN, NMA. Validation: CSN, AR, BR, PR, ÅL, TH, LK, NMA. Formal analysis: CSN, AR, BR, PR, AN, MC. Investigation: AR, BR, PR, ILF, MRP, SA, ÅL, TH, MS, LK, NMA. Resources: PR, ILF, MRP, SA, ÅL, CM. Data Curation: AR, TH, AN, LK, NMA. Funding acquisition: CSN, ÅL. Project administration: CSN, AR, PR. Supervision: CSN, PR, ÅL. Writing – original draft: CSN. Writing – review & editing: CSN, AR, BR, PR, ILF, MRP, AS, SA, AN, MS, ÅL, TH, MO, BS, CM, MC, LK, NMA

## Declaration of Interests

BS, AS have submitted a patent application for a COVID-19 peptide vaccine. Other authors declare that the research was conducted in the absence of any commercial or financial relationships that could be construed as a potential conflict of interest.

## Funding

This research was partially funded by the Swedish Research Council (VR: 2019-01736, 2017-05807, 2018-02569), The European Union’s Horizon 2020 Research Innovation Program under grant 874735 (VEO), The Knut and Alice Wallenberg Foundation, and the Science for Life Laboratory Uppsala (Projects: Nevermore Covid, SiCoV and Molres).

## Acknowledgments

We thank the SciLifeLab Autoimmunity and Serology Profiling infrastructure unit for multiplex bead array serology and Cecilia Hellström, Peter Nilsson and Sophia Hober for their valuable input. We thank Benjamin Murrell and Daniel Scheward for performing an initial pseudo neutralization assay for the project and for their valuable input in discussions of our data. We thank Jonna Hermansson and Lisa Lindberg for their help with sample collection at the elderly care home, Koon Chu Yaiw for sample preparations, Angela Silveira for providing the cohort of blood samples from 1996 and all individuals who donated blood to this study. Finally, we thank Stephen Ordway for excellent help with editing the manuscript.

## Data Sharing Statement

The datasets generated in current study are available from the corresponding authors upon reasonable request.

## Figure legends

**Supplementary Figure. 1**

**NGVEGF peptide specific antibodies are induced in mice by Flu or SARS-CoV-2 peptide vaccination**. (a and b) Mice received one dose of VaxigripTetra Quadrivalent vaccine (Sanofi Pasteur) followed 2 weeks later by a booster containing the same vaccine and 5 SARS-CoV-2 peptides (Table S2). Control mice were injected with saline. Sera were collected at 2 weeks (response after first dose) and at 6 weeks (response after second dose) and were analyzed for IgM (a) and IgG (b). OD values for IgM and IgG in control mice were <0.2. OD values for IgG after first dose vaccination was <0.2 and increase after second dose vaccination to >0.2. OD values for IgM was >0.2 at first dose vaccination and increased to >2.0. Nonparametric one-way ANOVA test and Dunn`s multiple comparison test were used. OD: optical density, ****p<0.00001, ***p<0.0001, **p<0.001, *p<0.01, ns: no significant.

## Notes

### Competing Interest Statement

BS and AS have submitted a patent application concerning a peptide Covid19 vaccine.

### Author Declarations

The study was approved by the Ethics Committee at Karolinska Institutet, Stockholm, Sweden (Dnr 2020-06333: all vaccinated subjects gave written informed consent; blood donors from blood bank were anonymous; Dnr 2020-07232: all subjects gave written informed consent; Dnr 06400: included anonymous blood samples from healthy donors (HD) in 2011 (Dnr: 01-420) and in 1996 (Dnr: 95-397 and 02-091). Ethical approval was obtained for the animal study (Dnr 16.07.2021).

### Summary of Updates

The manuscript has been reformatted for submission to another journal and include some minor updates of data analyses.

## References

1. N. M. Ferguson Dl, G. Nedjati-Gilani, N. Imai, K. Ainslie, M. Baguelin, S. Bhatia, A. Boonyasiri, Z. Cucunubá, G. Cuomo-Dannenburg, A. Dighe, I. Dorigatti, H. Fu, K. Gaythorpe, W. Green, A. Hamlet, W. Hinsley, L. C. Okell, S. van Elsland, H. Thompson, R. Verity, E. Volz, H. Wang, Y. Wang, P. G.T. Walker, C. Walters, P. Winskill, C. Whittaker, C. A Donnelly, S. Riley, A. C. Ghani. Report 9: Impact of non-pharmaceutical interventions (NPIs) to reduce COVID-19 mortality and healthcare demand. Imperial College London,. 2020;10(77482):491–7.

2. Huang C, Wang Y, Li X, Ren L, Zhao J, Hu Y, et al. Clinical features of patients infected with 2019 novel coronavirus in Wuhan, China. Lancet. 2020;395(10223):497–506.

3. Zhou F, Yu T, Du R, Fan G, Liu Y, Liu Z, et al. Clinical course and risk factors for mortality of adult inpatients with COVID-19 in Wuhan, China: a retrospective cohort study. Lancet. 2020;395(10229):1054–62.

4. O’Driscoll M, Dos Santos GR, Wang L, Cummings DAT, Azman AS, Paireau J, et al. Age-specific mortality and immunity patterns of SARS-CoV-2. Nature. 2020.

5. Moriarty LF, Plucinski MM, Marston BJ, Kurbatova EV, Knust B, Murray EL, et al. Public Health Responses to COVID-19 Outbreaks on Cruise Ships - Worldwide, February-March 2020. MMWR Morb Mortal Wkly Rep. 2020;69(12):347–52.

6. Expert Taskforce for the C-CSO. Epidemiology of COVID-19 Outbreak on Cruise Ship Quarantined at Yokohama, Japan, February 2020. Emerg Infect Dis. 2020;26(11):2591–7.

7. Rudberg AS, Havervall S, Manberg A, Jernbom Falk A, Aguilera K, Ng H, et al. SARS-CoV-2 exposure, symptoms and seroprevalence in healthcare workers in Sweden. Nat Commun. 2020;11(1):5064.

8. Madewell ZJ, Yang Y, Longini IM, Jr., Halloran ME, Dean NE. Household Transmission of SARS-CoV-2: A Systematic Review and Meta-analysis. JAMA Netw Open. 2020;3(12):e2031756.

9. Lindahl JF, Hoffman T, Esmaeilzadeh M, Olsen B, Winter R, Amer S, et al. High seroprevalence of SARS-CoV-2 in elderly care employees in Sweden. Infect Ecol Epidemiol. 2020;10(1):1789036.

10. Pathela P, Crawley A, Weiss D, Maldin B, Cornell J, Purdin J, et al. Seroprevalence of Severe Acute Respiratory Syndrome Coronavirus 2 Following the Largest Initial Epidemic Wave in the United States: Findings From New York City, 13 May to 21 July 2020. J Infect Dis. 2021;224(2):196–206.

11. Carlsson M, Hatem G, Söderberg-Nauclér C. Mathematical modeling suggests pre-existing immunity to SARS-CoV-2. MedRxiv. 2021.

12. Carlsson M, Söderberg-Nauclér C. Pre-immunity to influenza A H1N1 affects COVID-19 outbreak dynamics, predicts herd immunity thresholds, and implies that Stockholm has reached herd immunity twice. MedRxiv. 2021.

13. Grifoni A, Weiskopf D, Ramirez SI, Mateus J, Dan JM, Moderbacher CR, et al. Targets of T Cell Responses to SARS-CoV-2 Coronavirus in Humans with COVID-19 Disease and Unexposed Individuals. Cell. 2020;181(7):1489–501 e15.

14. Braun J, Loyal L, Frentsch M, Wendisch D, Georg P, Kurth F, et al. SARS-CoV-2-reactive T cells in healthy donors and patients with COVID-19. Nature. 2020;587(7833):270–4.

15. Le Bert N, Tan AT, Kunasegaran K, Tham CYL, Hafezi M, Chia A, et al. SARS-CoV-2-specific T cell immunity in cases of COVID-19 and SARS, and uninfected controls. Nature. 2020;584(7821):457–62.

16. Swadling L, Diniz MO, Schmidt NM, Amin OE, Chandran A, Shaw E, et al. Pre-existing polymerase-specific T cells expand in abortive seronegative SARS-CoV-2. Nature. 2021.

17. Sekine T, Perez-Potti A, Rivera-Ballesteros O, Stralin K, Gorin JB, Olsson A, et al. Robust T Cell Immunity in Convalescent Individuals with Asymptomatic or Mild COVID-19. Cell. 2020;183(1):158–68 e14.

18. Ma Z, Li P, Ikram A, Pan Q. Does Cross-neutralization of SARS-CoV-2 Only Relate to High Pathogenic Coronaviruses? Trends Immunol. 2020;41(10):851–3.

19. Jiang S, Du L. Effect of Low-Pathogenic Human Coronavirus-Specific Antibodies on SARS-CoV-2. Trends Immunol. 2020;41(10):853–4.

20. Ng KW, Faulkner N, Cornish GH, Rosa A, Harvey R, Hussain S, et al. Preexisting and de novo humoral immunity to SARS-CoV-2 in humans. Science. 2020.

21. Doshi P. Covid-19: Do many people have pre-existing immunity? BMJ. 2020;370:m3563.

22. Sette A, Crotty S. Pre-existing immunity to SARS-CoV-2: the knowns and unknowns. Nat Rev Immunol. 2020;20(8):457–8.

23. Lipsitch M, Grad YH, Sette A, Crotty S. Cross-reactive memory T cells and herd immunity to SARS-CoV-2. Nat Rev Immunol. 2020;20(11):709–13.

24. Greenhalgh T, Jimenez JL, Prather KA, Tufekci Z, Fisman D, Schooley R. Ten scientific reasons in support of airborne transmission of SARS-CoV-2. Lancet. 2021;397(10285):1603–5.

25. Bertoglio F, Meier D, Langreder N, Steinke S, Rand U, Simonelli L, et al. SARS-CoV-2 neutralizing human recombinant antibodies selected from pre-pandemic healthy donors binding at RBD-ACE2 interface. Nat Commun. 2021;12(1):1577.

26. Anderson EM, Goodwin EC, Verma A, Arevalo CP, Bolton MJ, Weirick ME, et al. Seasonal human coronavirus antibodies are boosted upon SARS-CoV-2 infection but not associated with protection. Cell. 2021;184(7):1858–64 e10.

27. Zhang Y, Aevermann BD, Anderson TK, Burke DF, Dauphin G, Gu Z, et al. Influenza Research Database: An integrated bioinformatics resource for influenza virus research. Nucleic Acids Res. 2017;45(D1):D466–D74.

28. Reynisson B, Alvarez B, Paul S, Peters B, Nielsen M. NetMHCpan-4.1 and NetMHCIIpan-4.0: improved predictions of MHC antigen presentation by concurrent motif deconvolution and integration of MS MHC eluted ligand data. Nucleic Acids Res. 2020;48(W1):W449–W54.

29. Ayoglu B, Mitsios N, Kockum I, Khademi M, Zandian A, Sjoberg R, et al. Anoctamin 2 identified as an autoimmune target in multiple sclerosis. Proc Natl Acad Sci U S A. 2016;113(8):2188–93.

30. Hober S, Hellstrom C, Olofsson J, Andersson E, Bergstrom S, Jernbom Falk A, et al. Systematic evaluation of SARS-CoV-2 antigens enables a highly specific and sensitive multiplex serological COVID-19 assay. Clin Transl Immunology. 2021;10(7):e1312.

31. Team, R.C.R. A language and environment for stastistical computing. R Foundation for Stastistical Computing, Vienna, Austria URL https://www.R-projectorg/. 2019.

32. Team, Studio RR. Integrated Development for R. RStudio, Inc, Boston MA URL http://www.rstudiocom/. 2018.

33. Rahbar A, Peredo I, Solberg NW, Taher C, Dzabic M, Xu X, et al. Discordant humoral and cellular immune responses to (CMV) in glioblastoma patients whose tumors are positive for CMV. Oncoimmunology. 2015;4(2):e982391.

34. Hoffman T, Kolstad L, Lindahl JF, Albinsson B, Bergqvist A, Ronnberg B, et al. Diagnostic Potential of a Luminex-Based Coronavirus Disease 2019 Suspension Immunoassay (COVID-19 SIA) for the Detection of Antibodies against SARS-CoV-2. Viruses. 2021;13(6).

35. Cori A, Ferguson NM, Fraser C, Cauchemez S. A new framework and software to estimate time-varying reproduction numbers during epidemics. Am J Epidemiol. 2013;178(9):1505–12.

36. Bi Q, Wu Y, Mei S, Ye C, Zou X, Zhang Z, et al. Epidemiology and transmission of COVID-19 in 391 cases and 1286 of their close contacts in Shenzhen, China: a retrospective cohort study. Lancet Infect Dis. 2020;20(8):911–9.

37. Yang J, Petitjean SJL, Koehler M, Zhang Q, Dumitru AC, Chen W, et al. Molecular interaction and inhibition of SARS-CoV-2 binding to the ACE2 receptor. Nat Commun. 2020;11(1):4541.

38. Liu WC, Lin CY, Tsou YT, Jan JT, Wu SC. Cross-Reactive Neuraminidase-Inhibiting Antibodies Elicited by Immunization with Recombinant Neuraminidase Proteins of H5N1 and Pandemic H1N1 Influenza A Viruses. J Virol. 2015;89(14):7224–34.

39. Gresset-Bourgeois V, Leventhal PS, Pepin S, Hollingsworth R, Kazek-Duret MP, De Bruijn I, et al. Quadrivalent inactivated influenza vaccine (VaxigripTetra). Expert Rev Vaccines. 2018;17(1):1–11.

40. Britton T, Ball F, Trapman P. A mathematical model reveals the influence of population heterogeneity on herd immunity to SARS-CoV-2. Science. 2020;369(6505):846–9.

41. Townsend JP, Hassler HB, Wang Z, Miura S, Singh J, Kumar S, et al. The durability of immunity against reinfection by SARS-CoV-2: a comparative evolutionary study. Lancet Microbe. 2021;2(12):e666–e75.

42. Verghese M, Jiang B, Iwai N, Mar M, Sahoo MK, Yamamoto F, et al. A SARS-CoV-2 Variant with L452R and E484Q Neutralization Resistance Mutations. J Clin Microbiol. 2021;59(7):e0074121.

43. Marin-Hernandez D, Schwartz RE, Nixon DF. Epidemiological evidence for association between higher influenza vaccine uptake in the elderly and lower COVID-19 deaths in Italy. J Med Virol. 2021;93(1):64–5.

44. Fink G, Orlova-Fink N, Schindler T, Grisi S, Ferrer APS, Daubenberger C, et al. Inactivated trivalent influenza vaccination is associated with lower mortality among patients with COVID-19 in Brazil. BMJ Evid Based Med. 2020.

45. Conlon A, Ashur C, Washer L, Eagle KA, Hofmann Bowman MA. Impact of the influenza vaccine on COVID-19 infection rates and severity. Am J Infect Control. 2021;49(6):694–700.

46. Debisarun PA, Gossling KL, Bulut O, Kilic G, Zoodsma M, Liu Z, et al. Induction of trained immunity by influenza vaccination - impact on COVID-19. PLoS Pathog. 2021;17(10):e1009928.

47. Jabr Alwazzeh M, Mohammed Telmesani L, Saud AlEnazi A, Abdulwahab Buohliqah L, Talal Halawani R, Jatoi NA, et al. Seasonal influenza vaccination coverage and its association with COVID-19 in Saudi Arabia. Inform Med Unlocked. 2021;27:100809.

48. Tayar E, Abdeen S, Abed Alah M, Chemaitelly H, Bougmiza I, Ayoub HK, AH., et al. ffectiveness of influenza vaccination against SARS-CoV-2 infection among healthcare workers in Qatar. medRxiv. 2022.

49. Rees-Spear C, Muir L, Griffith SA, Heaney J, Aldon Y, Snitselaar JL, et al. The effect of spike mutations on SARS-CoV-2 neutralization. Cell Rep. 2021;34(12):108890.

50. Gallagher KME, Leick MB, Larson RC, Berger TR, Katsis K, Yam JY, et al. SARS -CoV-2 T-cell immunity to variants of concern following vaccination. bioRxiv. 2021.

51. Graham C, Seow J, Huettner I, Khan H, Kouphou N, Acors S, et al. Impact of the B.1.1.7 variant on neutralizing monoclonal antibodies recognizing diverse epitopes on SARS-CoV-2 Spike. bioRxiv. 2021.

52. Liu Y, Liu J, Plante KS, Plante JA, Xie X, Zhang X, et al. The N501Y spike substitution enhances SARS-CoV-2 infection and transmission. Nature. 2021.

53. Khan A, Wei DQ, Kousar K, Abubaker J, Ahmad S, Ali J, et al. Preliminary Structural Data Revealed That the SARS-CoV-2 B.1.617 Variant’s RBD Binds to ACE2 Receptor Stronger Than the Wild Type to Enhance the Infectivity. Chembiochem. 2021;22(16):2641–9.

54. Wang R, Chen J, Gao K, Wei GW. Vaccine-escape and fast-growing mutations in the United Kingdom, the United States, Singapore, Spain, India, and other COVID-19-devastated countries. Genomics. 2021;113(4):2158–70.

55. Vasireddy D, Vanaparthy R, Mohan G, Malayala SV, Atluri P. Review of COVID-19 Variants and COVID-19 Vaccine Efficacy: What the Clinician Should Know? J Clin Med Res. 2021;13(6):317–25.

