## Supplementary figures and images for "Influenza A H1N1–mediated pre-existing immunity to SARS-CoV-2 predicts COVID-19 outbreak dynamics"

### Suppl Fig 1

#### Slide 1
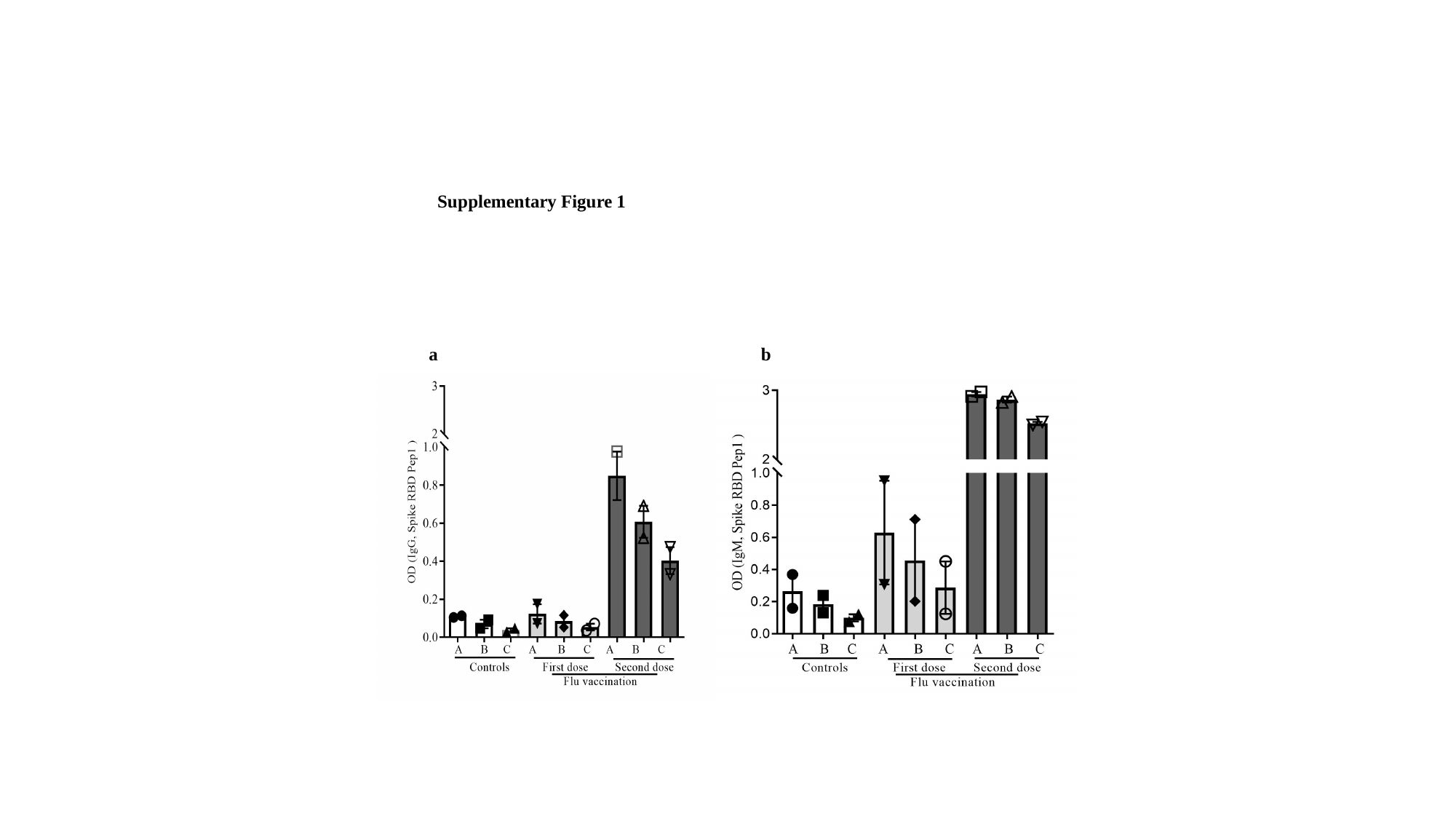
