## Supplementary text and tables for "Influenza A H1N1–mediated pre-existing immunity to SARS-CoV-2 predicts COVID-19 outbreak dynamics"

Supplementary Materials

Materials and Methods

Tables S1 to S4

References (*1-9*)

**Materials and Methods**

**Local BLAST search**

Local BLAST analyses were done with R (v3.5.3), RStudio (v1.1.463), and BLAST+ (v2.2.30+). PDB files were visualized with CCP4MG (v2.10.11). The local BLAST databases consisted of influenza protein retrieved from the Influenza Research Database. We then analyzed the SARS-CoV-2 spike protein reference strain (YP_009724390) by using a rolling window of 6 amino acids (6-mers). The 6-mers were blasted against the local influenza database with BLAST+ and the blastp routine in R. Sequences of 12 amino acid were further analyzed for potential HLA coverage and response. The IEDB MHC I binding prediction tool was used to analyze nine amino acids surrounding the identified 6-mers from the influenza proteins for their ability to bind to HLA class I molecules. All positive binders with corresponding HLA alleles for binders predicted to be of <1% percentile rank were selected. We then used HLA allele frequency maps (pypop.org) *(1)* to calculate the global and Scandinavian HLA coverage. One peptide was identical to a peptide, NGVEGF, in the neuraminidase of influenza A H1N1 Kyoto and Nagasaki strains. The MHCI binding predictions were made on 5/30/2021 with the IEDB analysis resource NetMHCpan (v4.1) tool *(2)*.

**ELISA**

Peptide-specific antibodies were detected as follows. Briefly, streptavidin (Sigma, CAT N° SA101, Sweden) diluted with phosphate buffered saline (PBS) was coated onto micro plates (Nunc AS, CAT N° 442404, Denmark) at a concentration of 10 ng per well and incubated for 2 h at 37°C. Wells were then coated with the spike peptide NGVEGF (H-PCNGVEGFNCYGGG(K(Biotin))-NH2, (Schafer-N), diluted in PBS with 1% bovine serum albumin (BSA) (Sigma, CAT N° 9418 Sweden) and 0.05% Tween-20 (Amresco, CAT N° 0777) to a concentration of 10 ng per well and incubated for 2 h at 37°C. Sera diluted 1:32 in PBS containing 1% BSA were added in duplicate and incubated overnight at 4°C. The plates were then incubated with alkaline phosphatase–conjugated anti-human IgG (1:3000, Merck, CAT N° AP113) for 1 h at 37°C and then with alkaline phosphatase substrate (Sigma, CAT N° P7998). The plates were washed three times with PBS and 0.05% Tween-20 after each incubation step. After 30 min of incubation at room temperature in the dark, the reaction was terminated by adding 3 mol/l NaOH (Substrate at Karolinska University Hospital). Absorbance at 405 nm (A405) was measured with a spectrophotometer (Versamax, Molecular Devices).

**Serology with a suspension bead array**

Protein and peptide bead arrays were prepared, and assays were done as described, with minor assay differences (*3*). Briefly, color-coded magnetic microspheres (MagPlex, Luminex) were covalently coupled to SARS-CoV-2 proteins (table S1); neutravidin (neutravidin 31000, Thermo Scientific) was used to couple the biotinylated peptides. All microspheres were pooled and mixed with the serum samples diluted 1:50 in assay buffer (PBS supplemented with 3% BSA, 5% non-fat milk, and 0.05% Tween-20). After incubation for 1 h, bound immunoglobulins were fixed with 0.2% paraformaldehyde (43368, Alfa Aesar) and incubated with anti-human IgG conjugated to R-phycoerythrin (H10104, Invitrogen; 12-4998-82, eBioscience) for 30 min, followed by detection of spike proteins with a FlexMap3D (Luminex Corp).

The suspension bead array data was processed using R (3.6.1) (*4*) in RStudio (1.2.1335) (*5*). Raw fluorescent intensities were used for the proteins while the signals for the peptides were transformed per sample into number of median absolute deviations (MADs) around the sample median to control for sample specific background levels. Seropositivity towards SARS-CoV-2 was determined as having reactivity towards two of three SARS-CoV-2 antigens (table S1) (*3*).

**Identification of IFN-γ−producing B and T cells by flow cytometry (FACS)**

Peripheral blood mononuclear cells (PBMCs) were isolated from blood samples with Lymphoprep (Stem Cell Technologies, CAT N° 7801) *(6)*, and reactivity of CD4 and CD8 T-cells and CD19 B-cells against the SPIKE 1 NGVEGF peptide (Schafer-N, Denmark) was analyzed by flow cytometry assay. Briefly, 5 × 10^4^ PBMCs were stimulated for 2 h with 1.6 μg of peptide and incubated overnight with brefeldin A (0.002 μg/μl; Sigma, CAT N° B6542) at 37°C in 5% CO_2_. Cells were washed with PBS (Gibco, CAT N° 14200067) and 1% BSA (Sigma, CAT N° 9418) and stained with mouse anti-human CD4 eFluor 450 (eBioscience, CAT N° 48-0048-41) and mouse anti-human CD8 PE (BD Biosciences, CAT N°555635) and mouse anti-human CD19 APC (BD Biosciences, CAT N°561742). After incubation for 30 min in the dark at 4°C, cells were fixed, permeabilized with Cell Fixation Cell Permeabilization Kit (Life Technologies, CAT N° GAS004) according to the manufacturer’s instructions), and stained with FITC-conjugated anti-interferon (IFN)-γ (BD Biosciences, CAT N° 561057) and analyzed by flow cytometry. Cells stimulated with staphylococcal enterotoxin B domain (SEB, Sigma, CAT N° S4881) served as positive controls. Negative cells were stained with corresponding isotypes: mouse anti-human IgG2b kappa eFluor 450 (eBiosciences, CAT N° 48-4732-80)) and mouse anti-human IgG1 kappa phycoerythrin and mouse anti-human-IgG1 kappa APC (both from BD Biosciences, CAT N° 555751). Cells were analyzed with a FACS machine (Novocyte, AH Diagnostics). The percentage of IFN-γ-positive cells in the isotype controls was subtracted from the specific stimulation.

**In vivo mouse model to test the NGVEGF peptide response after vaccination**

Wiss-Webster male mice aged 6–7 weeks were maintained under controlled conditions throughout the experiment (ambient temperature of 22 ± 2°C, a 12‑h light/dark cycle). The mice were housed 4 per cage and had free access to standard chow and water. The study protocol was approved by II Local Ethics Committee for Animal Experiments in Warsaw (Approval no. 16.07.2021). Experimental mice received the Vaxigrip vaccine (*n* = 12) on day 1; controls (*n* = 8) were not vaccinated. The vaccine was given on day 1 of the experiment, and a boost with the Vaxigrip vaccine coated with five SARS-CoV-2 peptides (CV-1–CV-5 (Biovacc-19) was given on day 23. Mice were anesthetized with isoflurane and decapitated, and whole blood was collected into sterile Eppendorf tubes 2 weeks after the first immunization (*n* = 6) and 4 weeks after the boost vaccination (*n* = 6). The samples were allowed to clot for 1 h at 37°C, cooled to 4°C, and centrifuged twice at 10,000 x *g* for 10 min each. Supernatants (serum) were aspirated and stored at 4°C.

*Vaccine preparation*: For the prime vaccination, VaxigripTetra Quadrivalent Flu vaccine (Sanofi Pasteur) was thawed, diluted 1:199 in saline, and 50 µl was injected subcutaneously in the neck with an insulin syringe; controls were injected with 50 µl of saline. Mice were randomly assigned to the groups. For boosting, a solution of 500 µl of Vaxigip, 50 µl of CV-1–CV-5 (Immunor; table S1), and 50 µl of saline was prepared and incubated at 4ºC for 3 days. On day 4, the solution was kept at room temperature for 8 h and frozen at –80ºC for 24 h. On the day of injection, the vaccine solution was thawed, diluted 1:143 in saline, and immediately injected at a dose of 50 µl.

**COVID-19 suspension immunoassay** (**SIA)**

The COVID-19 suspension immunoassay (SIA) was performed as described in (*7*). Briefly, the recombinant spike 1 (S1) antigen (40591-V08H, amino acids (aa) 16-685, Sino Biological, Beijing, China) was coupled to 2.5 × 10^6^ carboxylated differentially color-marked magnetic beads (MagPlex microspheres, Luminex Corp., Austin, Texas, US) using sulfo-N-hydroxysulfosuccinimide (sulfo-NHS) (ThermoFisher Scientific, Waltham, MA, USA) and 1-ethyl-3-[3 dimethylaminopropyl]carbodiimide hydrochloride (EDC) (Sigma Aldrich, Merck, Darmstedt, Germany), according to the manufacturer’s instructions. For SARS-CoV-2 specific IgG determination, serum diluted 1:25 (2 μl serum and 48 μl buffer) in PBSTT (phosphate-buffered saline supplemented with 0.5% Tween 20 and Tris (50 mM)) was added to 96-well microtiter plates. Vortexed and sonicated microsphere mixture (50 μl, 25 beads/μl PBSTT) was added to each well, giving a final serum dilution of 1:50. Subsequently, the plate was incubated for 60 min in the dark at room temperature on a plate shaker (400 rpm). Microspheres were then washed with 100 μl PBS, followed by addition of 100 μl (2 μg/ml PBSTT) biotinylated protein G (Pierce Biotechnology, ThermoFisher Scientific, Waltham, MA, USA), 30 min incubation, and washing. One hundred microliters (2 μg/ml PBSTT) streptavidin–phycoerythrin (SA-PE) (Invitrogen, ThermoFisher Scientific, Waltham, MA, USA) was then added, followed by an incubation period of 15 min. Finally, the microspheres were washed once before re-suspension in 100 μl PBS and analysis of 50 μl in a Luminex MagPix instrument (Luminex Corp., Austin, TX, USA). The assay cut-off for positivity was calculated as the average median fluorescence intensity (MFI) plus 6 SD plus 10% of 200 SARS-CoV-2 antibody-negative sera.

**SARS-CoV-2 surrogate virus neutralization test (sVNT)**

Recombinant SARS-CoV-2 S1 protein (40591-V08H, amino acids (aa) 16-685, Sino Biological, Beijing, China) was coupled to 2.5 x 10^6^ carboxylated differentially color-marked magnetic beads (MagPlex microspheres, Luminex) as described above for the COVID-19 SIA. Twenty-five microliter of biotinylated human angiotensin converting enzyme 2 (hACE2) (20 ng; 10108-H02H-B, Sino Biological, Beijing, China) and 5 μl serum sample (in 20 μl PBSTT) were incubated for 45 min with 50 μl vortexed and sonicated SARS-CoV-2 S1-coupled beads (25 beads/μl PBSTT), giving a final serum dilution of 1/20. Subsequently, the suspension was washed with 100 μl PBS, followed by addition of 100 μl (2 μg/ml in PBSTT) SA-PE (Invitrogen, ThermoFisher Scientific, Waltham, MA, USA), a 15 min incubation, another PBS wash, addition of 100 μl PBS, and briefly mixing the final reactions on a plate shaker. The MFI was determined as described above for the COVID-19 SIA. All incubations were performed in the dark at room temperature and on a plate shaker (400 rpm). PBSTT served as a negative control. Percentage binding inhibition (% BI) for each sample was calculated using the following formula:

% BI = (1 – (sample MFI/negative control MFI)) x 100.

**Mathematical modeling**

A simple mathematical test to estimate pre-immunity and R_0_ directly from a time series of cases (or deaths) was implemented, given that the seroprevalence is known both before and after the wave, and that NPIs do not change drastically during the elapse of the wave. Of critical importance for the computation of R_0_ is the assumption that everyone is susceptible. If we suppose that a fraction θ were immune at the onset of the pandemic, we arrive at the equation R_0,apparent_ = R_0,true_(1 – θ). As the pandemic unfolds, the effective R value, denoted R_e_ at time t depends only on s(t), the fraction of susceptible individuals in the population, and can be estimated with the simple formula

R_e(t)_ = R_0,true_s(t)	(1)

Note that s(0) = 1 – θ, so this is consistent with the previous formula for R_0_,apparent. Use of this formula to estimate the level of pre-existing immunity θ is based on a simple mathematical idea. Given a time series of cases I(t), we can estimate the right-hand side of (1) using any estimator of Re, (such as e.g. EpiEstim). In addition, we need a measured increase in sero-prevalence σ, by which we can estimate s(t) as

s(t) = (1-θ-ζ)- σ Iacc(t)/N,	(2)

where Iacc is the cumulative time series of cases and N is the total population, formed by summing the time series of new infections, which we denote by I. Here, θ is the pre-pandemic immunity and ζ is the immunity acquired before t = 0, which in the case of Stockholm was 0.1 at the onset of the second wave. With this information, (1) turns into a vector equation that can be solve by the least-squares method for the two unknowns R_0_ and θ. We set Θ = 100 θ (i.e., the percentage estimated to have been pre-immune at the onset of the pandemic).

**Discrepancy between new cases and daily PCR-positive tests**

We let ν(t) denote the time series of daily PCR-positive samples, averaged over a 7-day period. Ignoring the time between infection and testing positive and taking into account that only a fraction of positive cases gets tested, we arrive at

I(t) ≈ Cν(t),	(3)

where C is some unknown constant.

**Estimating R_e_**

To estimate R_e_, we used the online tool EpiEstim (https://shiny.dide.imperial.ac.uk/epiestim/) (*8*) and an estimate of the probability distribution of the serial interval for SARS-CoV-2 obtained from (*9*).

**Estimating i and s**

To estimate the function I – the fraction of infectious in SEIR, we used the time series ν for positive PCR tests (or deaths) and a point estimate of the seroprevalence σ. If we had access to I(t), then the total number of individuals who have or have had COVID-19 would equal Iacc(T) = ∑_(t≤T)▒〖I(t). In reality, we have access only to ν(t), so we introduce νacc(T) = ∑_(t≤T)▒〖ν(t)〗 which according to the approximation (2) satisfies Iacc = Cνacc. To accurately estimate the curve s(t) from a given population, we need to rely on serological studies. We will be interested in data from places where a serological study has determined the proportion Pτ of people having suffered COVID-19 at a given time τ. Formula (2) then becomes

s(t) = (1 − θ) −Pτ νacc(t) /νacc(τ) = (1 θ − ζ ) − ∆s(t),	(4)

where ∆s = Pτ νacc(t) /νacc(τ) measures the decrease in s from the onset of the pandemic.

**Estimating pre-immunity**

Since ∆s and Re are readily computable by data, this gives us the equation

R_e_(t) = R_0, true_(1 – θ − ζ) − R_0,true_∆s(t)_,_	(5)

which is easily solved with the least-squares method in the two unknowns R0, true and θ. However, it is important to realize that this formula is only valid while passing through the main part of infections. As cases decrease R_e_ (t) will be less than 1, but after the “wave” is over, crude macro-approximations such as (1) cease to be applicable. The left hand side of (6) will eventually oscillate around 1, whereas the right hand side of (6) is a decreasing function of time that will converge to some number <1. If there is no pre-immunity, then equation (6) has only one unknown variable R_0,true_, which again can be estimated by the least-squares method (Fig. 4). Applying this method to data from the second wave in Stockholm yields a pre-immunity of 60% (Fig. 4). In we assume no pre-immunity, the data fit is very poor, indicating that this is an unrealistic assumption.

**References**

1.	B. Reynisson, B. Alvarez, S. Paul, B. Peters, M. Nielsen, NetMHCpan-4.1 and NetMHCIIpan-4.0: improved predictions of MHC antigen presentation by concurrent motif deconvolution and integration of MS MHC eluted ligand data. *Nucleic Acids Res* **48**, W449-W454 (2020).

2.	B. Ayoglu *et al.*, Anoctamin 2 identified as an autoimmune target in multiple sclerosis. *Proc Natl Acad Sci U S A* **113**, 2188-2193 (2016).

3.	S. Hober *et al.*, Systematic evaluation of SARS-CoV-2 antigens enables a highly specific and sensitive multiplex serological COVID-19 assay. *Clin Transl Immunology* **10**, e1312 (2021).

4.	R Core Team (2019). R: A language and environment for statistical computing. R Foundation for Statistical Computing, Vienna, Austria. URL <https://www.R-project.org/>.

5.	Team, R. R. Studio, Integrated Development for R. *RStudio, Inc., Boston MA*, (2018). <http://www.rstudio.com/>.

6.	A. Rahbar *et al.*, Discordant humoral and cellular immune responses to (CMV) in glioblastoma patients whose tumors are positive for CMV. *Oncoimmunology* **4**, e982391 (2015)

7.	T. Hoffman *et al.*, Diagnostic Potential of a Luminex-Based Coronavirus Disease 2019 Suspension Immunoassay (COVID-19 SIA) for the Detection of Antibodies against SARS-CoV-2. *Viruses* **13**, (2021).

8.	A. Cori, N. M. Ferguson, C. Fraser, S. Cauchemez, A new framework and software to estimate time-varying reproduction numbers during epidemics. *Am J Epidemiol* **178**, 1505-1512 (2013).

9.	Q. Bi *et al.*, Epidemiology and transmission of COVID-19 in 391 cases and 1286 of their close contacts in Shenzhen, China: a retrospective cohort study. *Lancet Infect Dis* **20**, 911-919 (2020).

**Table S1**. Information about SARS-CoV-2 protein and peptide sequences used in the study.

| **Plot name** | **Sequence** |
| --- | --- |
| Spike S1S2 foldon  (Spike-f, Full length, *HEK*) | MFVFLVLLPLVSSQCVNLTTRTQLPPAYTNSFTRGVYYPDKVFRSSVLHSTQDLFLPFFSNVTWFHAIHVSGTNGTKRFDNPVLPFNDGVYFASTEKSNIIRGWIFGTTLDSKTQSLLIVNNATNVVIKVCEFQFCNDPFLGVYYHKNNKSWMESEFRVYSSANNCTFEYVSQPFLMDLEGKQGNFKNLREFVFKNIDGYFKIYSKHTPINLVRDLPQGFSALEPLVDLPIGINITRFQTLLALHRSYLTPGDSSSGWTAGAAAYYVGYLQPRTFLLKYNENGTITDAVDCALDPLSETKCTLKSFTVEKGIYQTSNFRVQPTESIVRFPNITNLCPFGEVFNATRFASVYAWNRKRISNCVADYSVLYNSASFSTFKCYGVSPTKLNDLCFTNVYADSFVIRGDEVRQIAPGQTGKIADYNYKLPDDFTGCVIAWNSNNLDSKVGGNYNYLYRLFRKSNLKPFERDISTEIYQAGSTPCNGVEGFNCYFPLQSYGFQPTNGVGYQPYRVVVLSFELLHAPATVCGPKKSTNLVKNKCVNFNFNGLTGTGVLTESNKKFLPFQQFGRDIADTTDAVRDPQTLEILDITPCSFGGVSVITPGTNTSNQVAVLYQDVNCTEVPVAIHADQLTPTWRVYSTGSNVFQTRAGCLIGAEHVNNSYECDIPIGAGICASYQTQTNSPGSASSVASQSIIAYTMSLGAENSVAYSNNSIAIPTNFTISVTTEILPVSMTKTSVDCTMYICGDSTECSNLLLQYGSFCTQLNRALTGIAVEQDKNTQEVFAQVKQIYKTPPIKDFGGFNFSQILPDPSKPSKRSFIEDLLFNKVTLADAGFIKQYGDCLGDIAARDLICAQKFNGLTVLPPLLTDEMIAQYTSALLAGTITSGWTFGAGAALQIPFAMQMAYRFNGIGVTQNVLYENQKLIANQFNSAIGKIQDSLSSTASALGKLQDVVNQNAQALNTLVKQLSSNFGAISSVLNDILSRLDPPEAEVQIDRLITGRLQSLQTYVTQQLIRAAEIRASANLAATKMSECVLGQSKRVDFCGKGYHLMSFPQSAPHGVVFLHVTYVPAQEKNFTTAPAICHDGKAHFPREGVFVSNGTHWFVTQRNFYEPQIITTDNTFVSGNCDVVIGIVNNTVYDPLQPELDSFKEELDKYFKNHTSPDVDLGDISGINASVVNIQKEIDRLNEVAKNLNESLIDLQELGKYEQGSGYIPEAPRDGQAYVRKDGEWVLLSTFLGRSLEVLFQGPGHHHHHHHHSAWSHPQFEKGGGSGGGGSGGSAWSHPQFEK |
| Spike S1  (Full length, *CHO*) | VNLTTRTQLPPAYTNSFTRGVYYPDKVFRSSVLHSTQDLFLPFFSNVTWFHAIHVSGTNGTKRFDNPVLPFNDGVYFASTEKSNIIRGWIFGTTLDSKTQSLLIVNNATNVVIKVCEFQFCNDPFLGVYYHKNNKSWMESEFRVYSSANNCTFEYVSQPFLMDLEGKQGNFKNLREFVFKNIDGYFKIYSKHTPINLVRDLPQGFSALEPLVDLPIGINITRFQTLLALHRSYLTPGDSSSGWTAGAAAYYVGYLQPRTFLLKYNENGTITDAVDCALDPLSETKCTLKSFTVEKGIYQTSNFRVQPTESIVRFPNITNLCPFGEVFNATRFASVYAWNRKRISNCVADYSVLYNSASFSTFKCYGVSPTKLNDLCFTNVYADSFVIRGDEVRQIAPGQTGKIADYNYKLPDDFTGCVIAWNSNNLDSKVGGNYNYLYRLFRKSNLKPFERDISTEIYQAGSTPCNGVEGFNCYFPLQSYGFQPTNGVGYQPYRVVVLSFELLHAPATVCGPKKSTNLVKNKCVNFNFNGLTGTGVLTESNKKFLPFQQFGRDIADTTDAVRDPQTLEILDITPCSFGGVSVITPGTNTSNQVAVLYQDVNCTEVPVAIHADQLTPTWRVYSTGSNVFQTRAGCLIGAEHVNNSYECDIPIGAGICASY |
| Nucleocapsid C  (*E. coli*) | TKKSAAEASKKPRQKRTATKAYNVTQAFGRRGPEQTQGNFGDQELIRQGTDYKHWPQIAQFAPSASAFFGMSRIGMEVTPSGTWLTYTGAIKLDDKDPNFKDQVILLNKHIDAYKTFP |
| NegIle1 | IIIIIIIIIIIIIIIIIIII |
| Spike peptide 1 | TLDSKTQSLLIVNNATNVVI |
| Spike peptide 2 | HKNNKSWMESEFRVYSSANN |
| Spike peptide 3 | KRISNCVADYSVLYNSASFS |
| Spike peptide 4 | STEIYQAGSTPCNGVEGFNC |
| Spike peptide 5 | PATVCGPKKSTNLVKNKCVN |
| Spike peptide 6 | DEMIAQYTSALLAGTITSGW |
| Spike peptide 7 | KYFKNHTSPDVDLGDISGIN |
| Spike RBD pep1 | H-PCNGVEGFNCYGGG(K(Biotin))-NH2 |
| Spike RBD pep2 | Biotin-(Ahx)GGGPCNGVEGFNCY-NH2 |
| Spike RBD pep3 | H-PCNGVEGFNCYFPLQSYGGG(K(Biotin))-NH2 |
| Spike RBD pep4 | Biotin-(Ahx)GGGPCNGVEGFNCYFPLQSYG-NH2 |
| Spike RBD pep5 | H-PCNGVKGFNCYGGG(K(Biotin))-NH2 |
| Spike RBD pep6 | Biotin-(Ahx)GGGPCNGVKGFNCYG-NH2 |
| Spike RBD pep7 | H-PCNGVKGFNCYFPLQSYGGG(K(Biotin))-NH2 |
| Spike RBD pep8 | Biotin-(Ahx)GGGPCNGVKGFNCYFPLQSYG-NH2 |
| Spike RBD pep9 | H-FRKSNLKPFERDISTGGG(K(Biotin))-NH2 |
| Spike RBD pep10 | Biotin-(Ahx)GGGFRKSNLKPFERDIST-NH2 |
| Spike RBD pep11 | H-ETIHNEEGVDWGGG(K(Biotin))-NH2 |
| Spike RBD pep12 | Biotin-GGGETIHNEEGVDW-NH2 |
| Biovacc-19 Peptide CV1 | RRGFKSYGVSPTKLNDSKVGGNYQNRLDSKVGGNY |
| Biovacc-19 Peptide CV2 | RRSTPSNGVERRGVEGFNENRFQPTNGRNRGVGYQP |
| Biovacc-19 Peptide CV3 | RRGASTEKSNRNGINITRQLLHAPATVRTNGVGYG |
| Biovacc-19 Peptide CV4 | RRKSTNLVGGQLTPTWGGGVKNKSVGGPLSETK |
| Biovacc-19 Peptide CV5 | RRFYPRGQGVFLEGATNTASWFRSRGFQFPRGQGIG |

**Table S2.** Frequency of HLA coverage of 12 Flu epitopes with SARS-CoV-2 similarity and allele frequency.

| Allele | Global frequency | Scandinavian frequency |
| --- | --- | --- |
| HLA-A*02:03 | 1.5% | 1.1% |
| HLA-A*02:06 | 3.5% | 1.3% |
| HLA-A*01:01 | 4.8% | 12.7% |
| HLA-A*26:01 | 3.4% | 3.2% |
| HLA-A*02:01 | 15.3% | 40.8% |
| HLA-A*30:02 | 1.5% | 0.8% |
| HLA-A*32:01 | 1.4% | 2.9% |
| HLA-A*33:01 | 1.2% | 0.6% |
| HLA-A*68:01 | 2.3% | 2.4% |
| HLA-A*31:01 | 4.1% | 3.6% |
| HLA-A*68:02 | 1.3% | 1.3% |
| Total | 40.2% | 70.7% |

Estimated global allele frequency is based on data from http://pypop.org.

**Table S3.** Flu peptides similar to SARS-CoV-2.

| Uniprot reference | Sequence |
| --- | --- |
| F8VBZ7 | KIVKLQDVVSIILTG |
| A0A2P1E2U3 | PVLYIDVADYSVDSS |
| A0A2U9I6I1 | LIVGISNLLLQVGNI |
| A0A455JC12 | PVLYVNVADYSVDSS |
| A0A190RUY6 | PVLYINVADYSVDSG |
| A0A2Z6FAF2 | ISYLLIVNNQNWSGN |
| A0A1B1PG85 | CIAWSSSSCHDGKAW |
| D0FHP1 | NDKHSNGTITDRSPY |
| A0A1D8DCN9 | NNGHSNNTVYDRTPY |
| A0A0S2RQT7 | NGANGVEGFSYRYGN |
| A0A190RVM8 | LYINVADYSVDAGYV |
| A0A5H2YHZ3 | PVLHINVADYSVDSG |

**Table S4.** Detection of anti-SARS-CoV-2 antibodies before vaccination, after flu vaccination and after COVID-19 vaccination using a COVID-19 suspension immunoassay (SIA) and a SARS-CoV-2 surrogate virus neutralization test (sVNT).

| Cohort | Age  (Year) | Sex | COVID-19 SIA (S1)  Cut-off = 300 MFI  IgG | | SARS-CoV-2 sVNT (S1)(S1)  All isotypes |
| --- | --- | --- | --- | --- | --- |
|  |  |  | MFI | Interpretation | BI (%) |
| 1 before vacc | >90 | M | 0 | | 21 |
| 1 after flu vacc |  |  | 0 | | 23 |
| 1 after SARS-CoV-2 vacc |  |  | 4490 P | | 79 |
| 2 before vacc | 80-90 | M | 0 | | 56 |
| 2 after flu vacc |  |  | 0 | | 32 |
| 2 after SARS-CoV-2 vacc |  |  | 2864 P | | 59 |
| 3 before vacc | 80-90 | F | 0 | | 24 |
| 3 after flu vacc |  |  | 1 | | 42 |
| 3 after SARS-CoV-2 vacc |  |  | 3617 P | | 65 |
| 4 before vacc | 70-90 | F | 0 | | 37 |
| 4 after flu vacc |  |  | 0 | | 44 |
| 4 after SARS-CoV-2 vacc |  |  | 3037 P | | 57 |
| 5 before vacc | 70-90 | F | 0 | | 34 |
| 5 after flu vacc |  |  | 0 | | 39 |
| 5 after SARS-CoV-2 vacc |  |  | 3347 P | | 61 |
| 6 before vacc | >90 | F | 0 | | 29 |
| 6 after flu vacc |  |  | 0 | | 34 |
| 6 after SARS-CoV-2 vacc |  |  | 567 P | | 42 |
| 7 before vacc | >90 | F | 0 | | 31 |
| 7 after flu vacc |  |  | 0 | | 45 |
| 7 after SARS-CoV-2 vacc |  |  | 6139 P | | 85 |
| 8 before vacc | 80-90 | F | 1363 P | | 52 |
| 8 after flu vacc |  |  | 1066 P | | 49 |
| 8 after SARS-CoV-2 vacc |  |  | 10800 P | | 99 |
| 9 before vacc | >90 | F | 0 | | 37 |
| 9 after flu vacc |  |  | 1 | | 47 |
| 9 after SARS-CoV-2 vacc |  |  | 759 P | | 34 |
| 10 before vacc | 80-90 | F | 0 | | 27 |
| 10 after flu vacc |  |  | 4 | | 32 |
| 10 after SARS-CoV-2 vacc |  |  | 1635 P | | 50 |
| 11 before vacc | 60-69 | M | 0 | | 32 |
| 11 after flu vacc |  |  | 0 | | 47 |
| 11 after SARS-CoV-2 vacc |  |  | 2857 P | | 60 |
| 12 before vacc | 80-89 | M | 0 | | 32 |
| 12 after flu vacc |  |  | 0 | | 48 |
| 12 after SARS-CoV-2 vacc |  |  | 23 | | 41 |
| 13 before vacc | 80-89 | F | 0 | | 50 |
| 13 after flu vacc |  |  | 0 | | 45 |
| 13 after SARS-CoV-2 vacc |  |  | 4517 P | | 84 |
| 14 before vacc | 70-79 | M | 0 | | 53 |
| 14 after flu vacc |  |  | 0 | | 47 |
| 14 after SARS-CoV-2 vacc |  |  | 9850 P | | 98 |
| 15 before vacc | 50-59 | M | 9 | | 47 |
| 15 after flu vacc |  |  | 0 | | 40 |
| 15 after SARS-CoV-2 vacc |  |  | 1762 P | | 59 |
| 16 before vacc | 20-29 | F | 0 | | 40 |
| 16 after flu vacc |  |  | 0 | | 45 |
| 16 after SARS-CoV-2 vacc |  |  | 1430 P | | 47 |
| 17 before vacc | 80-89 | M | 167 | | 38 |
| 17 after flu vacc |  |  | 244 | | 61 |
| 17 after SARS-CoV-2 vacc |  |  | 7598 P | | 98 |
| 18 before vacc | 70-79 | F | 0 | | 35 |
| 18 after flu vacc |  |  | 0 | | 56 |
| 18 after SARS-CoV-2 vacc |  |  | 8595 P | | 92 |
| 19 before vacc | 70-79 | M | 0 | | 25 |
| 19 after flu vacc |  |  | 0 | | 49 |
| 19 after SARS-CoV-2 vacc |  |  | 6611 P | | 92 |

M; male, F; female, P; positive, MFI; mean fluorescence intensity, vacc; vaccination, SIA; suspension immunoassay , S1; spike 1, sVNT; surrogate virus neutralization test, BI; binding inhibition
